# Comparison of ChatGPT vs. Bard to Anesthesia-related Queries

**DOI:** 10.1101/2023.06.29.23292057

**Authors:** Sourav S. Patnaik, Ulrike Hoffmann

## Abstract

We investigated the ability of large language models (LLMs) to answer anesthesia related queries prior to surgery from a patient’s point of view. In the study, we introduced textual data evaluation metrics, investigated “hallucinations” phenomenon, and evaluated feasibility of using LLMs at the patient-clinician interface. ChatGPT was found to be lengthier, intellectual, and effective in its response as compared to Bard. Upon clinical evaluation, no “hallucination” errors were reported from ChatGPT, whereas we observed a 30.3% error in response from Bard. ChatGPT responses were difficult to read (college level difficulty) while Bard responses were more conversational and about 8^th^ grade level from readability calculations. Linguistic quality of ChatGPT was found to be 19.7% greater for Bard (66.16 ± 13.42 vs. 55.27 ± 11.76; *p*=0.0037) and was independent of response length. Computational sentiment analysis revelated that polarity scores of on a Bard was significantly greater than ChatGPT (mean 0.16 vs. 0.11 on scale of −1 (negative) to 1 (positive); *p*=0.0323) and can be classified as “positive”; whereas subjectivity scores were similar across LLM’s (mean 0.54 vs 0.50 on a scale of 0 (objective) to 1 (subjective), *p*=0.3030). Even though the majority of the LLM responses were appropriate, at this stage these chatbots should be considered as a versatile clinical resource to assist communication between clinicians and patients, and not a replacement of essential pre-anesthesia consultation. Further efforts are needed to incorporate health literacy that will improve patient-clinical communications and ultimately, post-operative patient outcomes.

## Introduction

Pre-anesthesia evaluation is typically carried out 48-72 hours prior to the surgical procedure that requires administration of anesthesia. During this session, the anesthesiologists clarify numerous queries and misconceptions about anesthesia. Pre-operative instructions related to anesthesia can help alleviate post-operative recovery and can potentially help reduce patient’s anxiety, length of stay, and post-operative pain [1]. In fact, a systematic review found that majority of the discussion with patients have been focused on preoperative anesthesia planning, and not much details on postoperative critical care communications exist [2]. Hence, patient-anesthesiologist communication is critical for simple tasks such as imparting factual information and explaining simple anesthesia related concepts. Now the question is, can we use artificial intelligence tools to answer these critical pre-anesthesia questions?

We aimed to evaluate if the responses generated from popular large language models (LLMs), a type of Generative Artificial Intelligence that generates text-based contents, such as OpenAI ChatGPT [3] and Google Bard [4] would be comparable to a anesthesiologist’s response. Multidomain knowledge comprehension and problem-solving ability has been postulated as a specialty of theses LLM’s. Even though ChatGPT has aced several medical board examination questions [5–11] and was deemed more “empathetic” as compared to clinicians [12], some of the studies revealed that LLMs are not widely accepted by clinicians for consultation purposes and has received a lot of negative attention since their debut [13]. These advanced computing technologies have gained popularity due to their ease of use and applicability to cross-functional fields across the globe. LLMs like ChatGPT or Bard work best by a process of prompt engineering – a set of specific instructions that are provided to LLMs to elicit specific and relevant responses [14]. Based on the level of instructions provided, the prompts can be classified as *zero-shot* (minimal instructions are provided), *few-shot* (instructions are for input and output are provided with examples), and the more complex *chain-of-thought* (complex reasoning and detailed instructions are provided) [14, 15]. In this study, we focused on the *zero-shot* approach and evaluated the LLM responses with minimal instructions. Prior studies have similarly utilized zero-shot queries from patient’s perspective for other areas of medicine, [16–18] and primarily evaluated the AI generated responses through qualitative evaluations by clinicians. Our study aims to analyze the LLMs’ response to patient’s anesthesia-related queries in a more objective and quantitative manner. We have compared readability [16, 19–22], linguistic [23–26], and text-based analytical [13] differences between the two popular generative AI interfaces – ChatGPT and Bard.

## Methods

### Typical questions from patient’s prior during pre-anesthesia sessions

We selected commonly asked anesthesia related questions during the pre-anesthesia consultation procedure prior to surgery in a hospital setting. Table 1 provides the list of questions that were analyzed in this study:

**Table 1.**
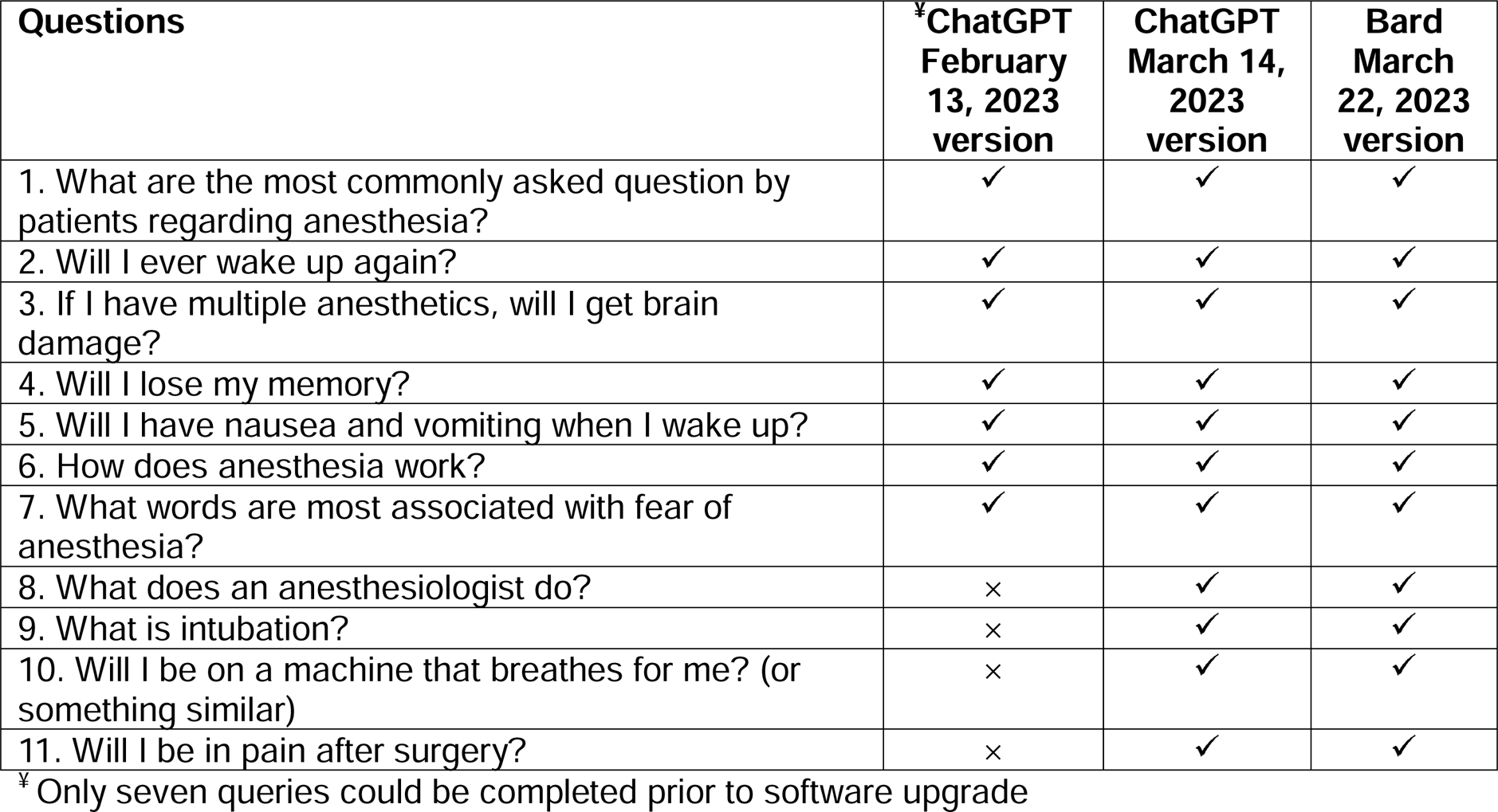
Anesthesia related questions commonly asked by patients during pre-anesthesia evaluation

| **Questions** | **\*ChatGPT**<br>**February**<br>**13, 2023**<br>**version** | **ChatGPT**<br>**March 14,**<br>**2023**<br>**version** | **Bard**<br>**March**<br>**22, 2023**<br>**version** |
| --- | --- | --- | --- |
| 1. What are the most commonly asked question by patients regarding anesthesia? | ✓ | ✓ | ✓ |
| 2. Will I ever wake up again? | ✓ | ✓ | ✓ |
| 3. If I have multiple anesthetics, will I get brain damage? | ✓ | ✓ | ✓ |
| 4. Will I lose my memory? | ✓ | ✓ | ✓ |
| 5. Will I have nausea and vomiting when I wake up? | ✓ | ✓ | ✓ |
| 6. How does anesthesia work? | ✓ | ✓ | ✓ |
| 7. What words are most associated with fear of anesthesia? | ✓ | ✓ | ✓ |
| 8. What does an anesthesiologist do? | × | ✓ | ✓ |
| 9. What is intubation? | × | ✓ | ✓ |
| 10. Will I be on a machine that breathes for me? (or something similar) | × | ✓ | ✓ |
| 11. Will I be in pain after surgery? | × | ✓ | ✓ |

\* Only seven queries could be completed prior to software upgrade

### Response generation from large language models

We used the ChatGPT February 13, 2023 version (ChatGPT(F)), or ChatGPT March 22, 2023 version (ChatGPT(M)) (OpenAI, San Francisco)[3] and experimental version of Google Bard March 23, 2023 version (Google, Mountain View, CA) [4] in this study. *Zero-shot* prompt were provided (minimal instructions to the LLMs [6, 27–30]) to the questions listed in Table 1. To have a standardized response across platforms/versions, we added a prefixed statement to each question – “*Focus on patients’ self-perceptions in preparation for anesthesia*”. The generated responses were saved in a .txt file, and further evaluated for quantitative analyses. These LLMs are constantly updated and hence, the results are interpreted based on the versions used on the day of assessment. All queries were repeated three times to reduce variability [31](total of 33 queries each for ChatGPT(M) (uses Generative Pre-trained Transformer architecture (GPT-3.5)) and Bard (uses LaMDA (Language Model for Dialogue Applications) architecture); only 7 queries for ChatGPT(F) due to limited access and software upgrades).

### Textual Response Evaluation

Detailed quantitative analysis for ChatGPT(F), ChatGPT(M), or Bard generated responses were evaluated using the metrics listed in Table 2.

**Table 2.** Metrics for analysis of responses generated by ChatGPT and Bard.

| Type of analysis | Metric | Interpretation | Score Ranges | Reference |
| --- | --- | --- | --- | --- |
| Reading assessment | Flesch Kincaid Grade Level (FKG) | Uses word length and sentence lengths to calculate how difficult the passage is to understand. E.g., a high score on the ease test should have a corresponding low score on the Grade-level test. | FKG score ranges from 0-20+; FRE ranges from 0 to 100 | [1-3] |
|  | Flesch-Kincaid readability reading ease (FKE) |  |  |  |
|  | SMOG score | Primarily used in healthcare. Focuses on polysyllabic words - words with multiple syllables | 0 to 211 or higher |  |
| Lexical diversity measures | Measure of textual lexical diversity (MTLD) | Measure of the range of different words in the text | Varies; lowest = 0 | [4, 5] |
| Computational Sentiment analysis | Polarity score | It indicates the sentiment polarity or sentiment orientation of a given text. It measures the degree of positivity, negativity, or neutrality in the text. | A normalized score of -1 to +1 is calculated for the text; 0 is designated as "neutral", between 0 to -1 is "negative", and between 0 to 1 is "positive" | [6, 7] |
|  | Subjectivity score | Subjectivity refers to the presence of personal opinions, beliefs, or emotions, while objectivity refers to the absence of personal bias or emotions, focusing more on factual information. | Subjectivity ranges from 0 to 1, with 0 being "objective" and anything >0.5 being "subjective" |  |
| Text similarity | Levenshtein distance | It's a measure of how different the two strings are or their linguistic distance. | Varies; lowest = 0 | [8, 9] |
| Others | Word counts (WC) | Total numbers of words in the text | lowest = 0 |  |
|  | Negative words count | Count of negative words associated with poor outcomes in pre-anesthesia consultation (-“aspiration”, “allergy”, “anaphylaxis, “local anesthetic”, “sedation”, “try”, and “worry” ) | lowest = 0 | [10, 11] |

### “Hallucinations”

“Hallucinations” are defined as errors in response from LLMs and these errors are framed by convincing statements that are not true. For this study, we are focusing on medical and surgery related facts. Factual accuracy of all LLM generated responses was qualitatively assessed by a clinician (UH). “Hallucination” error counts were expressed as a percentage of total responses (proportion).

### Readability assessment

Readability is the basic understanding of the text that is expressed in the form of mathematical formulas or indices. For each generated response, Flesch reading ease (FRE) (Eq.1), Flesch-Kincaid grade level scoring (FKG) (Eq.2), and Simple Measure of Gobbledygook (SMOG) (Eq.3) readability assessments were performed using the online resource - TextCompare.org [16, 32–34]. The quantification of textual readability is performed as follows:

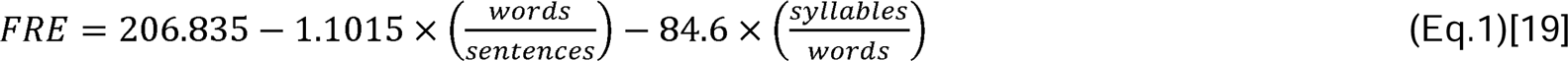

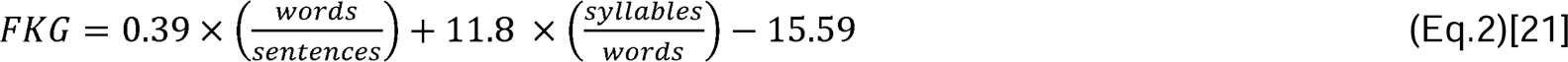

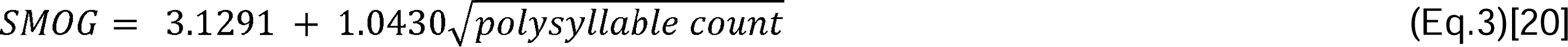

where polysyllable count is the number of words of three or more syllables per 30 sentences.

*FRE, FKG, and SMOG* scores were converted to grade equivalents as per table provided in Table S1(a-c) (supplementary materials). These assessments calculate how difficult the passage is to understand. For example, a high score on the Flesch ease test should have a corresponding low score on the Flesh-Kincaid grade-level test. Along the same lines, lower SMOG scores indicate high readability of the text. For patients, National Institutes of Health (NIH) recommends that the reading materials should be at a sixth grade reading level, whereas The American Medical Association (AMA) recommends an eight grade reading level [22, 35, 36]. Likewise, the NIH recommends the SMOG assessment for evaluating patient materials for clear and concise communication [20, 37]. Additionally, word count (WC) for each generated text was also calculated using the same online tool.

### Lexical Diversity measurements

The complexity of the text is defined here in terms of mathematical indicators named as measure of textual lexical diversity (MTLD). MTLD is defined as the mean length of sequential word strings that maintains its Type-Token Ratio (TTR) value (TTR is set to 0.72), where TTR is defined as a ratio of unique words (types) to the total number of words (token) in the text [23, 38]. We compared of the generated responses using an online MTLD tool developed by Reuneker et al. [26, 39]. Larger MTLD values indicate better linguistic capabilities and more complex writing abilities. In a clinical setting, MLTD measures have been previously utilized as biomarkers to distinguish written text from aphasia patients as compared to neurological intact ones [24].

### Computational Sentiment Analysis and Negative Word Detection

We performed computational sentiment analysis based on a vocabulary-based quantification or lexicon-based approach using *textblob 0.17.1* library (natural-language processing (NLP) tool) in *Jupyter Lab* environment (Python Interface) [40–42]. Words were extracted from the generated AI text and scored based on prior set rules or sentiment lexicon dictionary. Cumulative polarity and subjectivity scores were computed for each response. Polarity score between 0-1 was classified as “positive”, 0 was “neutral”, and less than zero until −1 was “negative”. Subjectivity score less than 0.5 were “objective”, and greater than 0.5 were “subjective”. For example, a positive score is given to words like “good”, “best”, “excellent”, etc., whereas negative score is given to words like “bad”, “awful”, “pathetic”, etc. Factual information constitutes lower subjectivity values and personal opinions constitute higher subjectivity values. In addition, we used the same Python package to scan the LLM generated texts for commonly misunderstood words during pre-anesthetic consultations - “aspiration”, “allergy”, “anaphylaxis, “local anesthetic”, “sedation”, “try”, and “worry” [43, 44].

### Effect of Iterations

Since each question was repeated three times, we wanted to evaluate if there were any variations in generated texts between ChatGPT and Bard. For this reason, we quantified the pairwise similarity between the generated texts were calculated using Levenshtein distance score via countwordsfree.com [45–48]. Levenshtein distance was introduced in 1966 [45] and has been adapted to measures how similar or dissimilar sentences are from each other. Larger distances mean sentences are very dissimilar from each other, and vice versa. Since ChatGPT(F) responses were available for only seven questions (without repetition), the comparison of the responses using Levenshtein distance was confined to ChatGPT(M) and Bard responses across the three iterations.

### Statistical Analysis

Unless stated otherwise, all data were expressed as mean ± std. deviation. Normality distribution of the data was checked using Shapiro Wilks test. All parameters, except for “Hallucinations” count, negative word count, and Levenshtein distance, were compared across the three LLMs (ChatGPT(F), ChatGPT(M), and Bard) using Kruskal-Wallis test and Dunn’s test for multiple comparisons. “Hallucination” and negative word counts were taken as proportion of whole count, and a z-score test of proportions was utilized to evaluate the differences between groups. Levenshtein distances between ChatGPT(M) and Bard were compared across three iterations (i.e., three repeats per query) using Kruskal-Wallis test and Dunn’s test for multiple comparisons. We wanted to evaluate if the length of the response influenced the quantified metrics from each LLM. We performed Spearman correlation analysis between all the quantified parameters vs. WC to evaluate the effect of number of words (WC) on each of the quantified parameters. We also evaluated the association between the polarity and objectivity scores generated from computational sentiment analysis using Spearman’s correlation analysis. All analyses were performed using GraphPad Prism ver 9.5.1 (GraphPad Software, San Diego, CA, USA,) and data was considered significant at *p<0.05*.

## Results

### Responses from LLMs

Representative screenshots of the response generated by ChatGPT and Bard to anesthesia related queries are shown in Figure 1.

**Fig. 1.**
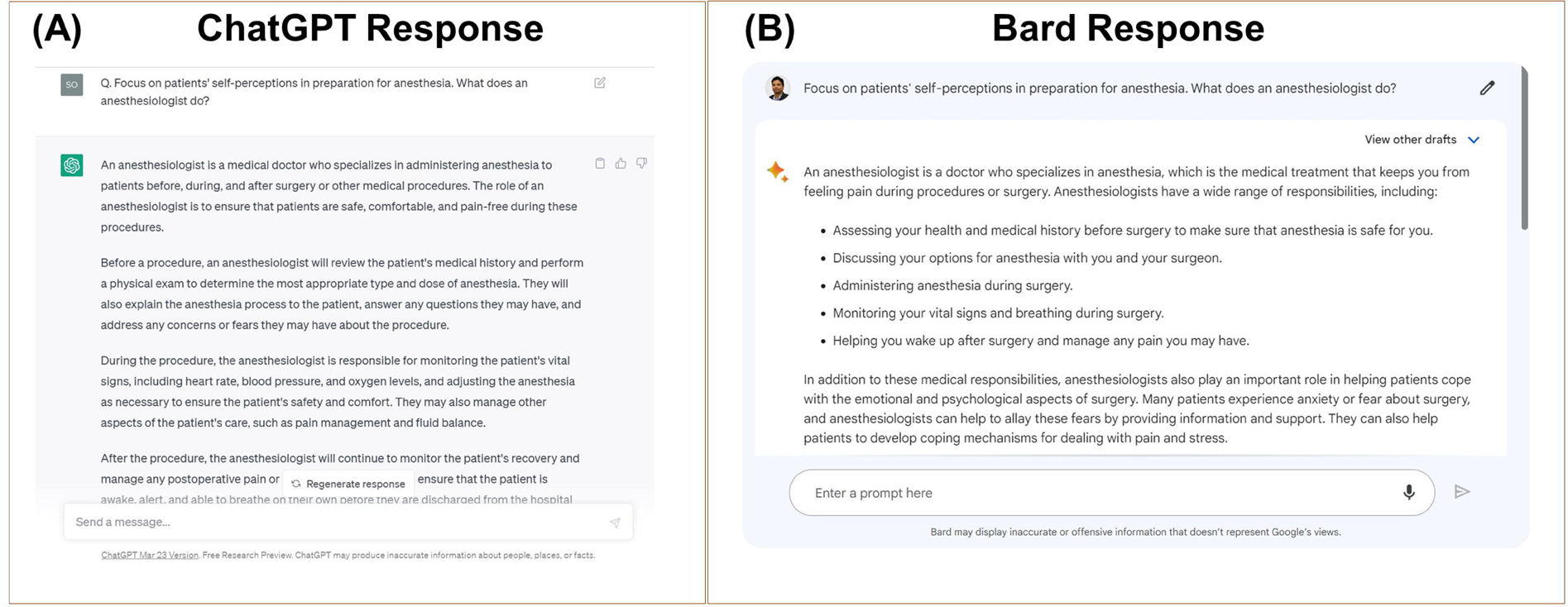
Exemplary response from ChatGPT (A) and Bard (B) to one of the patient queries related to anesthesia.

Detailed responses from all LLMs are provided in the S2(a-c) (Supplementary Materials). Qualitatively, responses from ChatGPT(F) or ChatGPT(M) were more refined, abstract, and adequate, as compared to Bard. Upon close examination, all the queries answered by ChatGPT versions were correct, whereas some of the Google Bard answers were incorrect (“hallucinations”) (Fig. 2; more details in S3 (supplementary materials)). Overall, accuracy of ChatGPT(F) (ChatGPT(F) vs. Bard - *z score* = 3.39; *p*=0.0007) and ChatGPT (M) (ChatGPT(M) vs. Bard - *z score* = 5.94; *p*<0.00001) were far superior (0/33; 0% error) to Google Bard (10/33; 30.3% error).

**Fig. 2.**
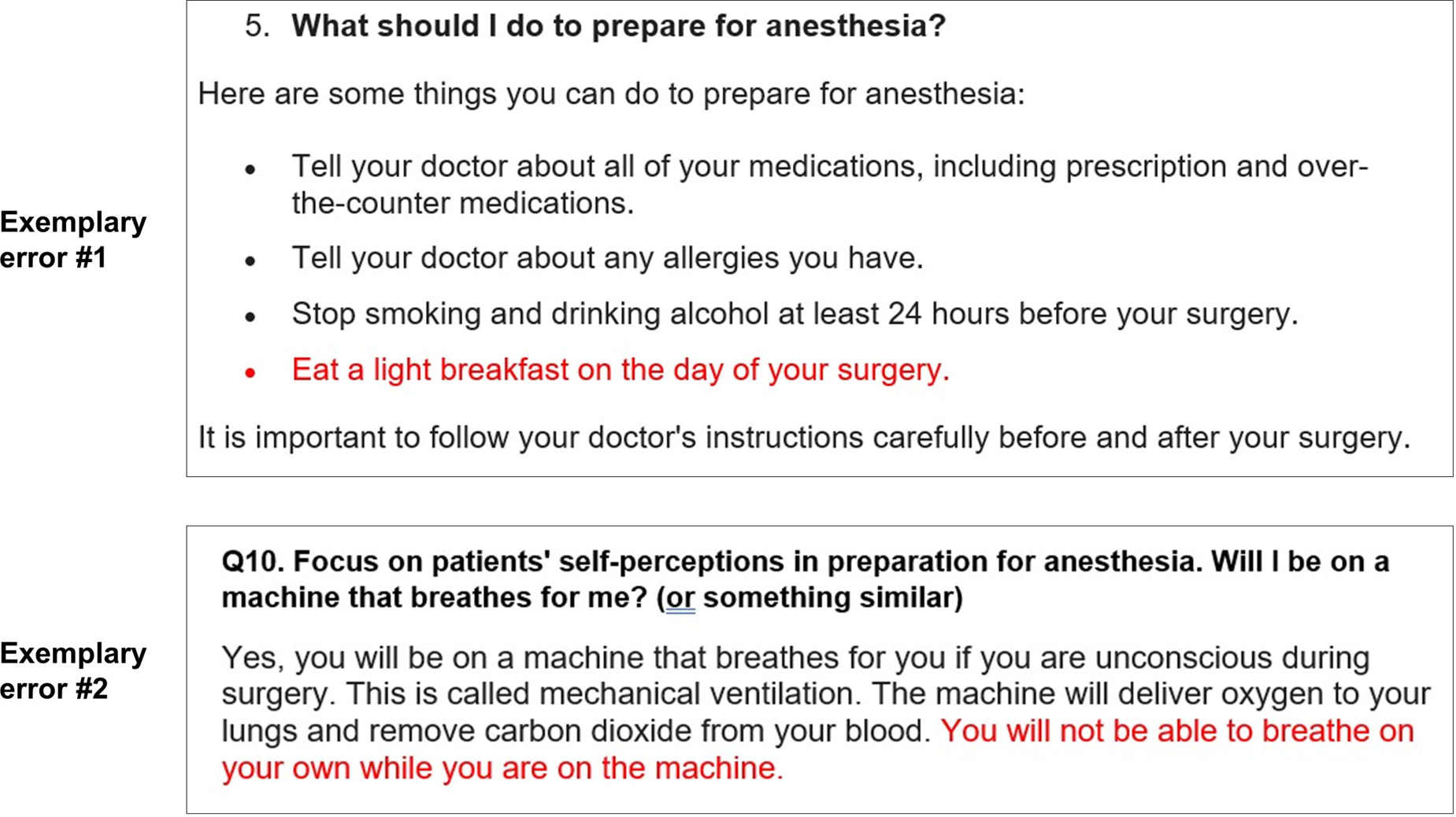
“Hallucinations” - exemplary errors from Google Bard in response to the anesthesia related queries.

### Readability assessment

FKG grade level scores were significantly higher for ChatGPT (M) as compared to ChatGPT (F) and Bard (14.74 ± 2.72 vs.14.36 ± 2.97 vs 9.4 ± 1.97, *p=*<0.0001) (Fig. 3(A)). With respect to the grade levels of ChatGPT(F), ChatGPT(M), and Bard correspond to “College level” (difficult to read), “College level” (difficult to read), and “8^th^ and 9^th^ grade” (conversational English), respectively (Table S1(a)). Similarly, FRE scores were found to 30 ± 21.21, 32.76 ± 14.14, and 55.35 ± 11.57 for ChatGPT(M), ChatGPT(F), and Bard, respectively (*p=*<0.0001) (Fig. 3(B)).

**Fig. 3.**
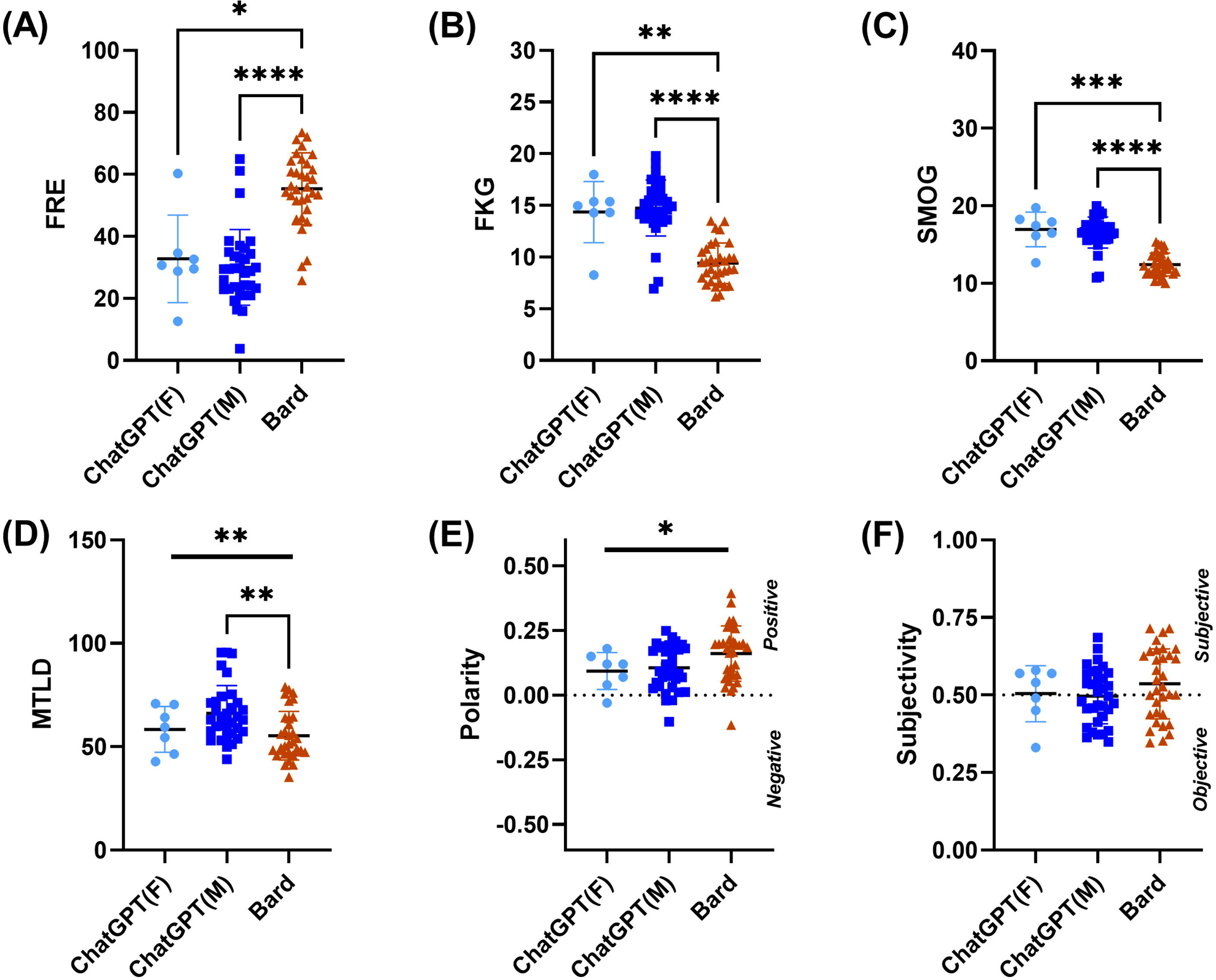
Detailed comparison of readability, linguistic, and text-based analytical of generated responses across ChatGPT(F), ChatGPT (M), and Bard. (A) FRG level, (B) FKE score, (C) SMOG, (D) MTLD, (E) Polarity, and (F) Objectivity parameters are compared across the three groups. Statistical significances are denoted by * (*p*<0.05), ** (*p*<0.005), **\*\*\* (*p*<0.001), and **** (*p*<0.0001).**

Conforming to Flesh-Kincaid Reading ease (Table (S1(b)), the FRE scores are at “College” levels for both ChatGPT versions, and “10th, 11th & 12th Grade” for Bard, respectively. SMOG scores of ChatGPT(F) and ChatGPT (M) were significantly different from Bard, but not statistically different amongst each other (16.94 ± 2.24 vs. 16.56 ± 2.0 vs.12.42 ± 1.49; *p* = <0.0001) (Fig. 3(C)). Per the SMOG index conversion (Table. S1(c)), the ChatGPT versions correspond to 7^th^ grade level (fairly easy to read) and Bard corresponds to 6th grade level (easy to read). In terms of word count (WC), Bard responses were shorter than ChatGPT(F) (167.1 ± 50.9 vs. 244.4 ± 34.72, *p* =0.0007) or ChatGPT (M) (167.1 ± 50.9 vs. 203.9 ± 37.64, *p* = 0.0052).

### Lexical diversity measures

MTLD quantification was found to be significantly different across the groups (*p =* 0.0037), but pairwise statistical differences were achieved only between ChatGPT(M) and Bard (66.16 ± 13.42 vs. 55.27 ± 11.76 vs. 58.35 ± 11.07) (Fig. 3(D)).

### Iteration effect

Levenshtein distances comparison across the three iterations of each question was similar for ChatGPT(M) and Bard (*p*=0.9335). Levenshtein distances between ChatGPT(M) and Bard were 1358.0 ± 253.6, 1341.0 ± 155.5, and 1356.0 ± 166.5 for first, second, and third iterations of the queries, respectively.

### Sentiment Analysis and Negative Words Detection

Polarity scores for Bard were significantly greater than the two ChatGPT versions (0.16 ± 0.11 vs. 0.09 ± 0.07 vs. 0.11 ± 0.09; *p*=0.0323) (Fig. 3(E)) and classified as “positive” for all LLMs. The subjectivity scores were similar across all three LLM’s – ChatGPT(F) 0.50 ± 0.09 vs. ChatGPT(M) 0.50 ± 0.09 vs. Bard 0.54 ± 0.11 (*p*=0.3030) (Fig. 3(F)) and classified as mildly “subjective”, even though statistically insignificant. The proportion of negative words for ChatGPT(F) were lower than ChatGPT(M) (0%(0/7) vs. 39.39% (13/33); z = −2.02, *p* = 0.04338); however, the proportion of negative words between ChatGPT(M) and Bard (39.39% (13/33) vs. 21.21% (7/33); z = 1.6, *p* = 0.1074), and ChatGPT(F) and Bard (0%(0/7) vs. 21.21% (7/33); z = −1.34, *p* = 0.1802), were similar.

### Effect of response length on quantified parameters

Association of word count with all other parameters are reported in Table 3. For ChatGPT(F), WC had no associations with any of the readability, lexical diversity, or sentiment analysis parameters. For ChatGPT(M), WC exhibited significantly moderate, negative association with polarity (ρ = −0.52; *p* = 0.0020) and subjectivity (ρ = −0.49; *p* = 0.0038) scores, respectively. Further, the polarity and subjectivity scores from ChatGPT(M) showed significant moderate, positive association with each other (ρ = 0.62; *p* = 0.0001). For Bard, WC exhibited significant, moderate negative association with MTLD only (ρ = −0.41; *p* = 0.0167).

**Table 3.** Association between variables quantified in this study. Bold numbers are significantly different p<0.05.

| Correlation Analysis |  | ChatGPT(F)<br>(n=7) |  | ChatGPT(M)<br>(n=33) |  | Bard<br>(n=33) |  |
| --- | --- | --- | --- | --- | --- | --- | --- |
| | | $\rho$ | *p-value* | $\rho$ | *p-value* | $\rho$ | *p-value* |
| WC | FKG | 0.59 | 0.1698 | 0.21 | 0.2415 | 0.15 | 0.4193 |
| WC | FKE | -0.57 | 0.2000 | -0.26 | 0.1399 | -0.28 | 0.1212 |
| WC | SMOG | 0.68 | 0.1095 | 0.20 | 0.2583 | 0.16 | 0.3713 |
| WC | MTLD | 0.29 | 0.5560 | -0.06 | 0.7274 | **-0.41** | **0.0167** |
| WC | Polarity | -0.29 | 0.5286 | **-0.52** | **0.0020** | -0.30 | 0.0860 |
| WC | Subjectivity | -0.56 | 0.2056 | **-0.49** | **0.0038** | -0.33 | 0.0591 |
| Polarity | Subjectivity | 0.18 | 0.6972 | **0.62** | **0.0001** | 0.26 | 0.1434 |

## Discussion

### LLMs in patient related queries

In this study, we (i) compared the error (“hallucination”) generated from LLMs when subjected to patient centric queries, (ii) showed that LLM’s are prompt dependent, (iii) introduced textual data assessment metrics for objective evaluation of LLM responses, and (iv) estimated the feasibility of LLM becoming a futuristic healthcare tool. We aimed to explore the LLM’s capability for future healthcare applications and more importantly if these AI-based chatbots are fit to answer patient’s queries accurately. Our goal was to perform a data analytics based quantitative assessment of the responses (from ChatGPT and Bard) to queries from patients’ point of view that were focused on anesthesia. Overall, ChatGPT has been more extensively tested for healthcare related and board exam questions as compared to Bard [5–8, 11, 16–18, 30, 34, 49–51]. To the best of our knowledge, this is the first study to quantitatively compare responses to anesthesia related questions between ChatGPT and Bard. Eleven common questions focusing on anesthesia were queried in chat interface of ChatGPT and Bard. The responses from these LLMs were then analyzed for word count, readability, linguistic quality (lexical diversity), sentiment analysis (polarity and subjectivity), and count of negative words commonly associated with poor surgical outcomes. A comparative account of the findings in this study and published literature are shown in Table 4.

**Table 4.**
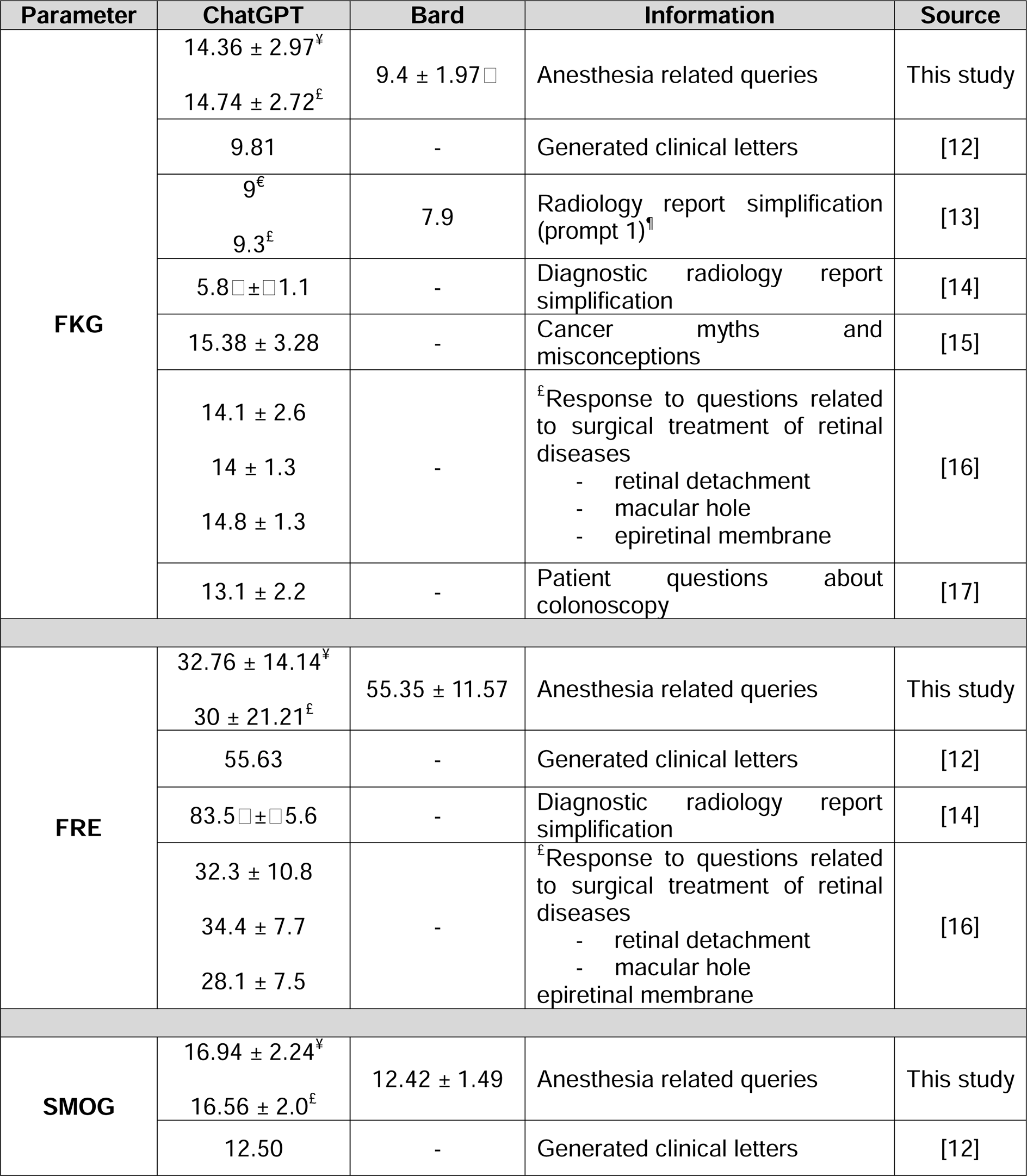
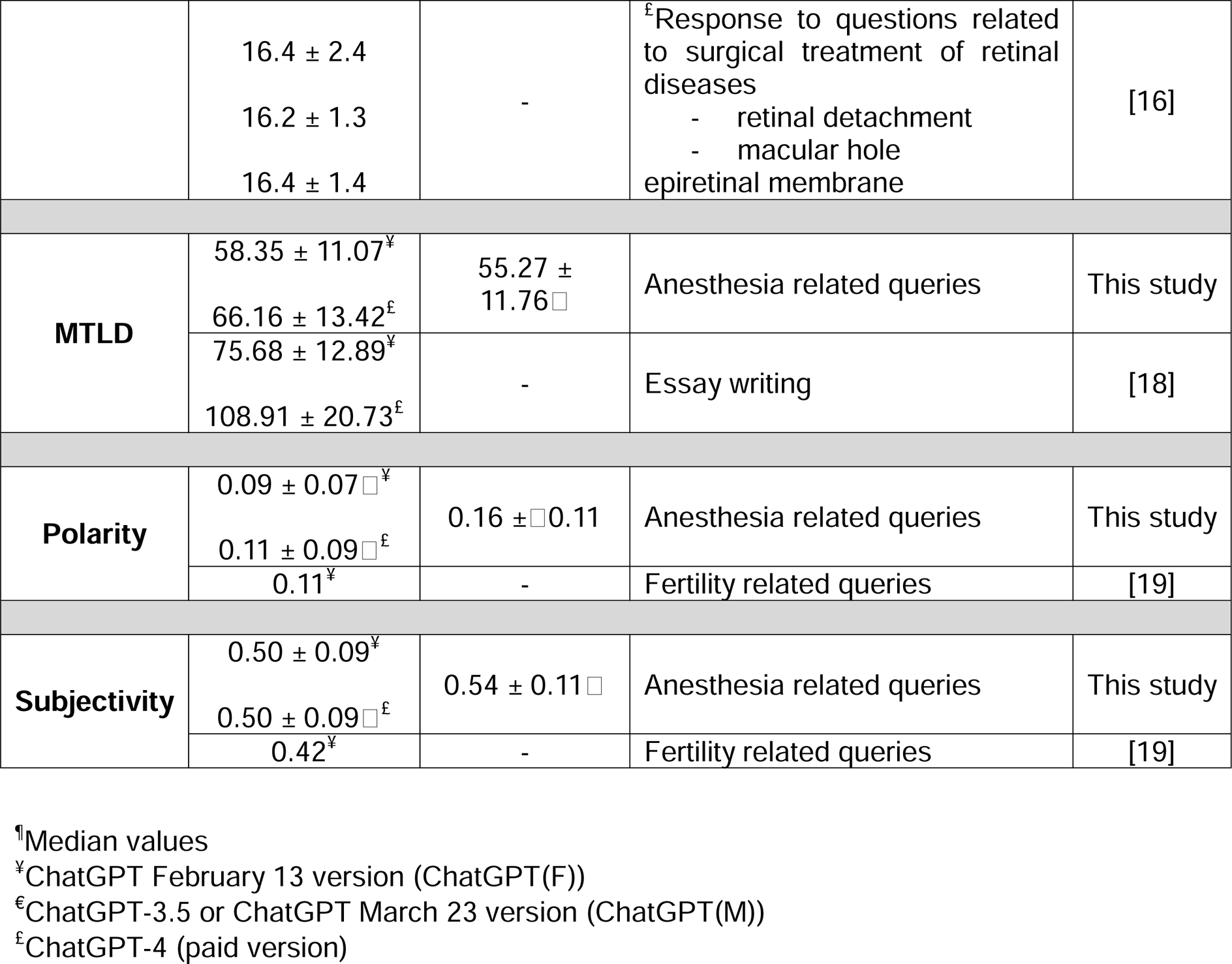
Comparative account of study parameters evaluated on ChatGPT and Bard responses

| Parameter | ChatGPT | Bard | Information | Source |
| --- | --- | --- | --- | --- |
| **FKG** | $14.36 \pm 2.97^{\text{¥}}$<br>$14.74 \pm 2.72^{\text{£}}$ | $9.4 \pm 1.97^{\square}$ | Anesthesia related queries | This study |
|  | 9.81 | - | Generated clinical letters | [12] |
| | $9^{\text{€}}$<br>$9.3^{\text{£}}$ | 7.9 | Radiology report simplification (prompt 1)<sup>¶</sup> | [13] |
| | $5.8^{\square} \pm^{\square} 1.1$ | - | Diagnostic radiology report simplification | [14] |
| | $15.38 \pm 3.28$ | - | Cancer myths and misconceptions | [15] |
| | $14.1 \pm 2.6$<br>$14 \pm 1.3$<br>$14.8 \pm 1.3$ | - | <sup>£</sup>Response to questions related to surgical treatment of retinal diseases<br>- retinal detachment<br>- macular hole<br>- epiretinal membrane | [16] |
| | $13.1 \pm 2.2$ | - | Patient questions about colonoscopy | [17] |
| **FRE** | $32.76 \pm 14.14^{\text{¥}}$<br>$30 \pm 21.21^{\text{£}}$ | $55.35 \pm 11.57$ | Anesthesia related queries | This study |
|  | 55.63 | - | Generated clinical letters | [12] |
| | $83.5^{\square} \pm^{\square} 5.6$ | - | Diagnostic radiology report simplification | [14] |
| | $32.3 \pm 10.8$<br>$34.4 \pm 7.7$<br>$28.1 \pm 7.5$ | - | <sup>£</sup>Response to questions related to surgical treatment of retinal diseases<br>- retinal detachment<br>- macular hole<br>epiretinal membrane | [16] |
| **SMOG** | $16.94 \pm 2.24^{\text{¥}}$<br>$16.56 \pm 2.0^{\text{£}}$ | $12.42 \pm 1.49$ | Anesthesia related queries | This study |
|  | 12.50 | - | Generated clinical letters | [12] |
|  | 16.4 ± 2.4<br>16.2 ± 1.3<br>16.4 ± 1.4 | - | <sup>£</sup>Response to questions related to surgical treatment of retinal diseases<br>- retinal detachment<br>- macular hole<br>epiretinal membrane | [16] |
| **MTLD** | 58.35 ± 11.07<sup>¥</sup> | 55.27 ± 11.76<sup>□</sup> | Anesthesia related queries | This study |
|  | 66.16 ± 13.42<sup>£</sup> |  |  |  |
|  | 75.68 ± 12.89<sup>¥</sup> | - | Essay writing | [18] |
|  | 108.91 ± 20.73<sup>£</sup> |  |  |  |
| **Polarity** | 0.09 ± 0.07<sup>□¥</sup> | 0.16 ± <sup>□</sup>0.11 | Anesthesia related queries | This study |
|  | 0.11 ± 0.09<sup>□£</sup> |  |  |  |
|  | 0.11<sup>¥</sup> | - | Fertility related queries | [19] |
| **Subjectivity** | 0.50 ± 0.09<sup>¥</sup> | 0.54 ± 0.11<sup>□</sup> | Anesthesia related queries | This study |
|  | 0.50 ± 0.09<sup>□£</sup> |  |  |  |
|  | 0.42<sup>¥</sup> | - | Fertility related queries | [19] |

<sup>¶</sup>Median values
<sup>¥</sup>ChatGPT February 13 version (ChatGPT(F))
<sup>€</sup>ChatGPT-3.5 or ChatGPT March 23 version (ChatGPT(M))
<sup>£</sup>ChatGPT-4 (paid version)

### ChatGPT outscored Bard in “Hallucinations”

Convincing, factually inaccurate responses with false claims (or “hallucinations”) is a common occurrence in LLM responses [56–59]. In the research arena, the most common artifact is fabricated references that clearly do not exist. One example would be the investigation by Grigio et al. [60] where they queried ChatGPT for the use of anti-emetic drug, Olanzapine, for postoperative nausea and vomiting. ChatGPT generated four references in support of the Olanzapine query; however, none of the references generated were found in research databases. To the best of our knowledge, this is the first study to report “hallucinations” from Bard in response to anesthesia related queries (Fig.2). We found that ChatGPT, overall, has the upper hand when it comes to responses to patient queries and did not provide any incorrect answers or hallucinations, as opposed to Bard. The potential application of large language models such as ChatGPT for answering presurgical anesthesia related questions was suggested by Bhattacharya et al. [56]; however, their focus was more surgical oriented and theoretical in nature. Angel et al.[7] compared the responses from GPT-3, GPT-4, and Bard from “Anesthesia Review: 1000 Questions and Answers to Blast the BASICS and Ace the ADVANCED” question bank and found that GPT-4 had the highest scores (78.33%) followed by GPT-3 (58.33%) and Bard (46.67%). Shay et al.[11] reported 56% success in ChatGPT responses to question from “Anesthesiology Examination and Board Review” book. Next, Aldridge et al. [5] reported moderate efficiency (63.6% accurate) of ChatGPT in answering Royal College of Anaesthetists fellowship exam questions (FRCA). Similarly, Birkett et al.[8] queried FRCA multiple choice questions and found that ChatGPT underperformed (proportion of correct responses was 0.697) and fell short of the official pass marks. Similar to our findings, a study comparing GPT-4, ChatGPT-3.5, and Bard performed on a neurosurgery oral boards preparatory question bank showed that Bard showed lowest performance score with accuracy of 44.2% (and highest “hallucination” rate), followed by ChatGPT-3.5 (62.4%) and GPT-4 (82.6%) [6]. Contrary to previous findings [5–7, 11], the LLM “hallucinations” was 0% for ChatGPT and 30.3% for Bard for anesthesia related queries. Concurring with previous studies [6, 7], we also report that responses from both ChatGPT versions were far superior and more accurate than Bard.

### Readability of Bard was better than ChatGPT

Bard was less “wordier” compared to both ChatGPT versions (167.1 ± 50.9 vs. 244.4 ± 34.72 vs. 203.9 ± 37.64), and its responses in our study were comparable to ChatGPT studies by Johnson et al. (172.62 ± 34.77) [16] and Lee et al. (175.1 ± 58.2) [54]. Readability analysis of Bard responses using FKG, FRE, and SMOG were found to be at “8^th^ and 9^th^ grade”, “10^th^, 11^th^ & 12^th^ Grade”, and “easy to read”, respectively (Table 4). For Bard, the FKG levels align with the AMA level requirements [22, 35, 36] and the SMOG score concurs with NIH level requirements [20, 37], but the FRE is slight greater than both established norms. For ChatGPT(F) and ChatGPT(M), the readability for FKG and FRE was “college level” and the SMOG scores were “fairly easy to read”. Like our study, Johnson et al. [16] addressed similar addressed cancer related myths and misconceptions using ChatGPT and found that there were noticeable higher FKG readability scores for ChatGPT (15.38 ± 3.28; “college level”) as compared to National Cancer Institute’s (NCI’s) answers (12.04 ± 2.42; 10^th^, 11^th^ & 12^th^ Grade”). The only other study comparing readability of ChatGPT and Bard responses was reported by Doshi et al. [49] and they investigated FKG score of radiological report simplification. Comparable to our study, they reported that Bard showed readability of “7^th^ grade level”, whereas ChatGPT showed readability of “8^th^-9^th^ grade level”. A study by Ali et al. [52] was the only one that compared the readability of three tools, FKG, FKE, and SMOG, for evaluating ChatGPT responses for generating clinical letters. Contrary to our investigation, they reported “8^th^-9^th^ grade level”, “10th, 11th & 12th Grade”, and “easy to read”, for FKG, FKE, and SMOG scores, respectively. Several other studies focusing on ChatGPT are either better [55] or similar [31, 54] readability scores as compared to our findings, are listed in Table 4. Differences in readability measures between ChatGPT and Bard stem from their original computational architecture. Bard is developed on LaMDA (Language Model for Dialogue Applications), trained on 137 billion parameters, and it fetches data from the internet for each query. Whereas ChatGPT is powered by Generative Pre-trained Transformer (GPT), trained on 175 billion parameters, and is designed to possess natural language understanding, reasoning ability, and can generate coherent text in response to prompts. Since Bard was designed as a service-oriented tool, it thus possesses the ability to use simple words and craft sentences that is easily “readable”.

### LLM’s are prompt dependent

Prompt engineering has opened up a wide arena for LLMs [15], and is considered as a backbone of many AI based tool today. Nastasi et al. [50] evaluated ChatGPT’s responses from patients’ perspective on acute illness, and queried critical questions concerning race, gender, and insurance status, and found the responses to be unsuited for personalized medical advice. They found ChatGPT changed clinical recommendations when social factors such as race or insurance status were modified. For this reason, we focused on a *zero-shot* learning approach that provides direct queries and minimal instructions to LLM to generate a response. Aldridge et al. [5] queried Royal College of Anaesthetists fellowship exam questions with GPT-3.5 and GPT-4, and found that ChatGPT(M) (utilizing GPT-4 technology) was able to answer questions with higher accuracy and exhibited a learning effect when subjected to repeated queries. To minimize variability, we repeated our queries three times, non-sequentially and found that pairwise Levenshtein distance scores to find similarities between the responses across the two LLMs (refer to *Iteration effect* in “Results” section). Thus, the generated responses from LLMs are user-centric, prompt-dependent, and their performance improves with repetition of queries.

### ChatGPT exhibited more lexical diversity than Bard

We introduced a linguistic analysis for evaluating responses from LLMs, which has been previously utilized as a biomarker for aphasia patients [23]. MTLD is a robust lexical diversity metric and has been shown to be impartial to word count for a range of vocabularies and word counts/tokens [25]. A study by Herbold et al.[53] performed MTLD based lexical diversity analysis of essays written by human authors, ChatGPT-3, and ChatGPT-4, and found that ChatGPT-4 has more complex and diverse vocabulary compared to human authors or ChatGPT3 (108.91 ± 20.73 vs. 95.72 ± 23.50 vs 75.68 ± 12.89, respectively). Along the same lines, both ChatGPT versions were not influenced by word counts, and were greater in lexical diversity than Bard. Furthermore, Bard showed a negative association with increasing word count, and this could be since Bard’s framework is designed to “conversational” and generates more human-like conversation, as compared to ChatGPT.

### Sentiment analysis and negative word detection

Sentiment analysis quantification is typically utilized for classifying public sentiment in social media platforms, reviews, etc. We applied the same technique to LLM responses and quantified the generated responses for polarity (positive, neutral, or negative) and objectivity (objective or subjective) score quantification. Close to Chervenak et al. [42], we found that there was no difference across ChatGPT and Bard in the overall objectivity scores – meaning that LLM responses are typically indifferent to queries. However, we did find that Bard was more “positive” as compared to the two ChatGPT versions. This is possibly since Bard utilized NLP techniques to stimulate “human” like conversations and on the other hand, ChatGPT is designed to be better at “summarization” of complicated healthcare data [30, 42, 49, 52, 55].

### Limitations and Future work

Variability in responses from ChatGPT and Bard across users must be addressed. Incomplete or inappropriate responses are quite common in LLM related queries. We repeated the questions three times and found no difference between the overall text outputs (Levenshtein distance metric). It is possible that repeating the queries can improve the overall performance of LLMs, as reported previously [5]. However, studies have utilized various number of iterations and there is no objective way to narrow down a specific iteration number that would work best for all scenarios. We have only utilized the free version of ChatGPT (GPT-3.5 based) in this study and did not find many differences in the quantified metrics between the two versions of ChatGPT as access to the older version was limited due to software upgrades. Further, it is possible that there may be differences in responses to the queries in the paid version of ChatGPT (GPT-4 based). As noted by Nastasi et al. [50], there were noted differences in LLM response when the prompt included patient socioeconomic conditions, health insurance status, etc. Hence, more studies should focus on patient based LLM responses to include patient social determinants of health in future studies. Differences in initial prompts (like the *prefixed statement* utilized in this study) can impact the overall performance and accuracy of ChatGPT responses [6]. To take a step further, future studies can focus on level of complexity of the queries by incorporating the *Chain-of-Thought Prompting* (a step-by-step reasoning prior to answering questions)[29], as compared to *zero shot* (text-based instructions – this study) and/or *few-shot* approaches (text-based instructions with examples of input-output provided)[27, 28]. In addition to the response evaluation methods used in this study, several text-based quantifications such as automated evaluation metrics (ROUGE-L, METEOR, etc.), relationship extraction, dependency parsing, and intent extraction can be explored as well [30, 61–63]. The “hallucination” phenomenon of ChatGPT is a well-known problem and has been reported by several publications [56–59], and in fact, we show the Bard showed “hallucinations” in its responses as well. There is always a risk of fabricated evidence [60] to support a “hallucination” by LLMs, and that is not acceptable when it comes to communication of facts to a patients. The most important question – does the LLM “understand” the query given by the user like a human does? That question is still not clear. The development of LLMs for future healthcare tools would require additional layers of scrutiny and comprehension before it can be used for mainstream healthcare practices.

## Conclusion

Our study shows that LLMs can generate effective responses from patient queries. ChatGPT was technical, precise, and descriptive, whereas Bard was conversational, adequate and exhibited “hallucinations”. The best utilization of LLMs in a patient centric scenario would be for generating text for effective patient communication prior to surgery (this study), summarizing concise radiological report [49, 55], and many others faceted improvements in quality of perianesthesia patient care [64, 65]. Restriction of LLMs is a mere stopgap solution and more efforts are needed to integrate with current technology for improvement of patient care and advancement of medical research. It is important to note that LLM’s cannot fully “think” and distinguish correction information from misinformation (yet). Creativity and ethical judgement are the hallmarks of a clinician, and that cannot be replaced with any technology.

## Data Availability

All data produced in the present work in provided in the supplementary materials.

