## Supplementary data for "Comparison of ChatGPT vs. Bard to Anesthesia-related Queries"

### Table S1. Interpretation of readability formulas

#### S1(a) Flesch-Kincaid Grade Level

| **Score** | **School Level** | **Comprehension** |
| --- | --- | --- |
| 5.0-5.9 | 5th Grade | Very easy to read |
| 6.0-6.9 | 6th Grade | Easy to read |
| 7.0-7.9 | 7th Grade | Fairly easy to read |
| 8.0-9.9 | 8th & 9th Grade | Conversational English |
| 10.0-12.9 | 10th , 11th & 12th Grade | Fairly difficult to read |
| 13.0-15.9 | College | Difficult to read |
| 16.0-17.9 | College Graduate | Very difficult to read |
| 18.0+ | Professional | Extremely difficult to read |

#### S1(b) Flesch Reading Ease

| **Score** | **School Level** | **Comprehension** |
| --- | --- | --- |
| 100.0-90.0 | 5th Grade | Very easy to read |
| 90.0-80.0 | 6th Grade | Easy to read |
| 80.0-70.0 | 7th Grade | Fairly easy to read |
| 70.0-60.0 | 8th & 9th Grade | Conversational English |
| 60.0-50.0 | 10th, 11th & 12th Grade | Fairly difficult to read |
| 50.0-30.0 | College | Difficult to read |
| 30.0-10.0 | College Graduate | Very difficult to read |
| 10.0-0.0 | Professional | Extremely difficult to read |

#### S1(c) SMOG Index

| **Total Number of Polysyllabic Words** | **School Level** | | **Comprehension** |
| --- | --- | --- | --- |
| 0-2 | 4th Grade | Very easy to read | |
| 3-6 | 5th Grade | Very easy to read | |
| 7-12 | 6th Grade | Easy to read | |
| 13-20 | 7th Grade | Fairly easy to read | |
| 21-42 | 8th & 9th Grade | Conversational English | |
| 43-56 | 10th Grade | Fairly difficult to read | |
| 57-72 | 11th Grade | Fairly difficult to read | |
| 73-90 | 12th Grade | Fairly difficult to read | |
| 91-110 | College Freshman | Difficult to read | |
| 111-132 | College Sophomore | Difficult to read | |
| 133-156 | College Junior | Difficult to read | |
| 157-182 | College Senior | Difficult to read | |
| 183-210 | College Graduate | Very difficult to read | |
| 211+ | Professional | Extremely difficult to read | |

### S2. Detailed responses to the questions from ChatGPT(F), ChatGPT(M), and Bard

#### S2 (a) ChatGPT(F) responses to the queries listed in Table 1 (Q.1-7)

**Q1. Focus on patients' self-perceptions preparation for anesthesia. What are the most commonly asked question by patients regarding anesthesia?**

Patients who are preparing for anesthesia may have various concerns and questions related to the procedure. Some of the most common questions that patients ask regarding anesthesia include:

1. What type of anesthesia will be used for my procedure?
2. How long will the anesthesia last?
3. What are the potential risks and complications associated with anesthesia?
4. How will the anesthesia be administered?
5. Will I be awake or asleep during the procedure?
6. How will I feel after the anesthesia wears off?
7. What can I expect during the recovery period?
8. Are there any steps I should take before the procedure to prepare for anesthesia?
9. Can I eat or drink anything before the procedure?
10. Can I take my regular medications before the procedure?

It's important for patients to have an open and honest conversation with their anesthesia provider to address any concerns or questions they may have. This can help patients feel more comfortable and confident about the anesthesia process and may help to reduce any anxiety or fear they may be experiencing.

**Q2. Focus on patients' self-perceptions preparation for anesthesia. Will I ever wake up again?**

It is understandable for patients to be concerned about the risks associated with anesthesia and whether they will wake up after the procedure. However, it's important to note that modern anesthesia is generally safe, and serious complications are rare.

Anesthesia providers are highly trained and take many precautions to ensure the safety of their patients. They carefully monitor patients' vital signs throughout the procedure, and the drugs used for anesthesia are carefully chosen and dosed based on the patient's individual needs.

While there is always a small risk of complications with any medical procedure, the vast majority of patients wake up after anesthesia without any problems. In rare cases where complications occur, anesthesia providers are trained to quickly identify and manage any issues that arise.

It's also important to note that patients who have pre-existing medical conditions, such as heart disease or lung disease, may have a higher risk of complications with anesthesia. Patients who have concerns about their individual risks should discuss these concerns with their anesthesia provider prior to the procedure.

Overall, while there is always some risk associated with anesthesia, the vast majority of patients undergo anesthesia safely and wake up without any issues. Patients who have concerns about anesthesia should discuss these concerns with their anesthesia provider prior to the procedure to help alleviate any fears or anxiety they may be experiencing.

**Q3. Focus on patients' self-perceptions in preparation for anesthesia. If I have multiple anesthetics, will I get brain damage?**

It is understandable for patients to have concerns about the potential risks of anesthesia, including the possibility of brain damage. However, it's important to note that the use of modern anesthesia is generally safe and brain damage is an extremely rare complication.

Anesthesia providers are highly trained to carefully monitor patients throughout the procedure to ensure their safety. They use advanced equipment to monitor vital signs such as blood pressure, heart rate, and oxygen levels to ensure that patients are stable and receiving the appropriate level of anesthesia.

While it is possible for some patients to experience complications related to anesthesia, such as allergic reactions or breathing difficulties, the risk of brain damage is extremely low. Multiple exposures to anesthesia, such as for multiple surgeries or procedures, do not increase the risk of brain damage or other serious complications.

That being said, patients who have pre-existing medical conditions, such as heart disease or lung disease, may have a higher risk of complications with anesthesia. Additionally, patients who have a history of alcohol or drug abuse or who have experienced brain injury in the past may have an increased risk of complications with anesthesia.

Overall, while there is always some risk associated with anesthesia, the use of modern anesthesia is generally safe and complications, including brain damage, are extremely rare. Patients who have concerns about their individual risks should discuss these concerns with their anesthesia provider prior to the procedure to help alleviate any fears or anxiety they may be experiencing.

**Q4. Focus on patients' self-perceptions in preparation for anesthesia. Will I lose my memory?**

It's common for patients to have concerns about the potential impact of anesthesia on their memory. While some patients may experience temporary memory loss or confusion after anesthesia, it is generally not a long-term or permanent issue.

The type of anesthesia used, the length of the procedure, and the individual patient's health and medical history can all play a role in the potential impact of anesthesia on memory. In general, however, the vast majority of patients do not experience long-term memory loss or other cognitive impairments as a result of anesthesia.

It's important to note that some patients may experience temporary confusion or memory loss after anesthesia, especially if they have had a long or complex procedure. This is typically a short-term issue and resolves within a few hours or days after the procedure.

Patients who have concerns about the potential impact of anesthesia on their memory should discuss these concerns with their anesthesia provider prior to the procedure. In some cases, an anesthesia provider may recommend a different type of anesthesia or adjust the dosing of the anesthesia to help minimize any potential impact on memory or cognitive function.

Overall, while some patients may experience temporary memory loss or confusion after anesthesia, the vast majority of patients do not experience long-term or permanent cognitive impairments as a result of anesthesia. Patients who have concerns about anesthesia should discuss these concerns with their anesthesia provider to help alleviate any fears or anxiety they may be experiencing.

**Q5. Focus on patients' self-perceptions in preparation for anesthesia. Will I have nausea and vomiting when I wake up?**

It is common for patients to experience nausea and vomiting after anesthesia. The exact cause of postoperative nausea and vomiting is not well understood, but it is believed to be related to a variety of factors such as the type and dose of anesthesia, the length of the procedure, and the individual patient's medical history.

To help prevent nausea and vomiting after anesthesia, anesthesia providers may administer medications before, during, or after the procedure. These medications may include anti-nausea medications or other drugs to help prevent or manage potential side effects.

However, despite these precautions, some patients may still experience nausea and vomiting after anesthesia. If you are concerned about nausea and vomiting after anesthesia, it's important to discuss these concerns with your anesthesia provider before the procedure. They may be able to adjust the medications or dosing to help minimize the risk of these side effects.

In addition, there are some steps that patients can take to help reduce the risk of nausea and vomiting after anesthesia, such as avoiding food or drink for a certain period of time before the procedure or gradually reintroducing food and drink after the procedure. Your anesthesia provider will provide specific instructions for your individual needs.

Overall, while it is common for patients to experience nausea and vomiting after anesthesia, there are steps that can be taken to help prevent or manage these side effects. Patients who have concerns about these side effects should discuss these concerns with their anesthesia provider to help alleviate any fears or anxiety they may be experiencing.

**Q6. Focus on patients' self-perceptions in preparation for anesthesia. How does anesthesia work?**

Anesthesia is a medical treatment that involves the administration of medications to produce a temporary loss of sensation and/or consciousness, allowing patients to undergo procedures without experiencing pain or discomfort. The exact mechanisms of how anesthesia works are complex and not yet fully understood, but it generally involves the following three main components:

1. Hypnosis: This component of anesthesia involves inducing a state of unconsciousness or reduced consciousness. This is typically achieved through the administration of intravenous or inhaled medications that act on the brain to produce sedation and unconsciousness.
2. Analgesia: This component of anesthesia involves reducing or eliminating pain sensation. This is typically achieved through the administration of local anesthetics, opioids, or other medications that act on the central nervous system to block pain signals.
3. Muscle relaxation: This component of anesthesia involves reducing muscle activity or inducing temporary paralysis. This is typically achieved through the administration of medications that act on the neuromuscular junction to block the transmission of nerve signals to muscles.

Together, these components of anesthesia work to produce a state of unconsciousness, reduce pain sensation, and induce muscle relaxation, allowing patients to undergo procedures without experiencing pain or discomfort.

The exact type of anesthesia used and the specific medications and dosing will vary depending on the individual patient's medical history, the type of procedure being performed, and other factors. Anesthesia providers are highly trained to assess each patient's individual needs and develop a customized anesthesia plan that is safe and effective for their specific situation.

Overall, while the exact mechanisms of how anesthesia works are complex and not yet fully understood, anesthesia is a safe and effective medical treatment that allows patients to undergo procedures without experiencing pain or discomfort.

**Q7. Focus on patients' self-perceptions in preparation for anesthesia. What words are most associated with fear of anesthesia?**

The fear of anesthesia can elicit a variety of emotions in patients, and there are several words and phrases that are commonly associated with this fear. Here are some of the most commonly associated words and phrases:

1. Loss of control: Many patients fear the loss of control that comes with anesthesia, especially if they are being put under completely.
2. Fear of the unknown: Patients may fear the unknown aspects of the anesthesia process, including the medications used, potential side effects, and the experience of being under anesthesia.
3. Awareness during anesthesia: Patients may fear the possibility of being aware or conscious during the procedure, despite being under anesthesia.
4. Side effects: Patients may fear potential side effects of anesthesia, including nausea and vomiting, dizziness, or headache.
5. Pain and discomfort: Some patients may fear experiencing pain or discomfort during the procedure, despite being under anesthesia.
6. Complications: Patients may fear potential complications or adverse events associated with anesthesia, including allergic reactions or breathing problems.

It's important to note that while these fears are understandable, anesthesia is a safe and routine medical treatment that is used to help patients undergo procedures without experiencing pain or discomfort. Anesthesia providers are highly trained to assess each patient's individual needs and develop a customized anesthesia plan that is safe and effective for their specific situation. If you have fears or concerns about anesthesia, it's important to discuss them with your anesthesia provider before the procedure. They can help alleviate any fears or anxiety you may be experiencing and provide you with more information about the anesthesia process.

#### S2 (b) ChatGPT(M) responses to the queries listed in Table 1 (Q.1-11)

**Q1.** **Focus on patients' self-perceptions preparation for anesthesia. What are the most commonly asked question by patients regarding anesthesia?**

When patients are preparing for anesthesia, they may have many questions about the process and what to expect. Some of the most commonly asked questions include:

- What type of anesthesia will I receive?
- How long will the anesthesia last?
- Will I be conscious during the procedure?
- What are the risks and side effects of anesthesia?
- How will I feel after the procedure?
- What can I do to prepare for anesthesia?
- What should I expect during the recovery period?
- Can I eat or drink before anesthesia?
- Will I be able to drive after the procedure?
- How will anesthesia affect my medical conditions or medications?

These are just a few of the many questions that patients may have about anesthesia. It's important for patients to ask their healthcare provider any questions they may have so they can feel informed and comfortable with the anesthesia process.

**Iteration 2** Retrieved on 04/27/2023

As an AI language model, I don't have access to current data or experiences of real patients. However, based on general knowledge and past research, some of the most common questions that patients ask about anesthesia include:

- What type of anesthesia will I receive?
- How will the anesthesia be administered?
- Will I be awake during the procedure?
- What are the risks and side effects of anesthesia?
- How long will the effects of anesthesia last?
- Will I experience pain after the procedure?
- How will I feel after waking up from anesthesia?
- Can I eat or drink before anesthesia?
- What should I do to prepare for anesthesia?
- How can I minimize the risk of complications during and after anesthesia?

It's important for patients to feel informed and comfortable about the anesthesia process. If you have any concerns or questions about anesthesia, it's best to discuss them with your healthcare provider before the procedure.

**Iteration 3** Retrieved on 04/27/2023

Patients may have a variety of questions and concerns about anesthesia, depending on their individual experiences and knowledge. Some of the most commonly asked questions by patients regarding anesthesia may include:

1. What type of anesthesia will be used for my procedure?
2. How will anesthesia affect me during and after the procedure?
3. Will I be conscious or asleep during the procedure?
4. What are the risks and potential complications of anesthesia?
5. How can I prepare for anesthesia, and are there any special instructions I should follow before my procedure?
6. Will I experience any pain or discomfort during the procedure, and if so, how will it be managed?
7. How long will the effects of the anesthesia last?
8. What kind of monitoring will be used during the procedure to ensure my safety?
9. How will anesthesia interact with any medications I am currently taking?
10. What can I expect during the recovery period, and how long will it take to fully recover from the effects of anesthesia?

It is important for patients to discuss any concerns or questions they may have with their healthcare provider, including their anesthesiologist, prior to their procedure to ensure they are fully informed and prepared.

Retrieved on 03/14/2023

**Q2. Focus on patients' self-perceptions preparation for anesthesia. Will I ever wake up again?**

It is completely normal for patients to have concerns and fears about waking up after anesthesia. However, it is important to note that the risk of not waking up after anesthesia is extremely low.

Anesthesia is a carefully monitored and controlled process, and anesthesiologists are highly trained professionals who take many precautions to ensure patient safety. The medications used during anesthesia are carefully titrated to achieve the desired level of sedation or unconsciousness while minimizing the risk of complications.

In addition, modern anesthesia machines are equipped with sophisticated monitoring devices that track the patient's vital signs, such as heart rate, blood pressure, and oxygen saturation. These machines also have safety features that can alert the anesthesiologist if there is a problem during the procedure.

It's important to have open and honest communication with your healthcare team about any concerns or fears you may have regarding anesthesia. They can help address your concerns and provide reassurance about the safety of the procedure.

**Iteration 2** Retrieved on 04/27/2023

It is natural for patients to feel anxious about the anesthesia process, and a common fear is whether they will wake up again after being put to sleep. However, it's important to understand that modern anesthesia is generally safe, and serious complications are rare.

Anesthesia medications are carefully selected and dosed based on a patient's individual needs, medical history, and the type of procedure being performed. Additionally, anesthesia is typically administered by a highly trained anesthesia provider who monitors the patient's vital signs and adjusts the anesthesia as needed throughout the procedure.

While there is always a small risk of complications with anesthesia, the vast majority of patients wake up from anesthesia without incident. Your healthcare provider can provide you with more specific information about your individual risks and how they will be mitigated during your procedure.

If you have concerns about waking up from anesthesia, it's important to discuss them with your healthcare provider before the procedure. They can provide you with information about the anesthesia process, answer any questions you may have, and help alleviate any anxiety you may be experiencing.

**Iteration 3** Retrieved on 04/27/2023

It is understandable for patients to have concerns about waking up from anesthesia, but it is important to understand that anesthesia is a highly controlled and monitored process. The anesthesiologist will be closely monitoring the patient's vital signs and adjusting the amount of medication administered to ensure that the patient remains unconscious throughout the procedure while also waking up safely afterwards.

It is also important to note that modern anesthesia techniques are generally very safe, and the risk of serious complications or death is very low. While every medical procedure carries some degree of risk, the benefits of anesthesia generally far outweigh the risks.

That being said, patients can take steps to help ensure a safe and successful anesthesia experience. This includes following any instructions provided by their healthcare provider, disclosing all medical conditions and medications to their anesthesiologist, and asking any questions or expressing any concerns they may have before the procedure.

In the rare event that a patient experiences complications during or after anesthesia, their healthcare team will be prepared to address any issues and provide appropriate care.

Retrieved on 03/14/2023

**Q3. Focus on patients' self-perceptions in preparation for anesthesia. If I have multiple anesthetics, will I get brain damage?**

It is understandable that patients may have concerns about the potential risks of multiple anesthesia exposures and the possibility of brain damage. However, it is important to note that the risk of brain damage from anesthesia is very low, even with multiple exposures.

The medications used during anesthesia are carefully titrated and monitored to achieve the desired level of sedation or unconsciousness while minimizing the risk of complications. Additionally, the anesthesiologist and healthcare team closely monitor the patient's vital signs, including brain function, during the procedure.

Research studies have shown that there is no evidence that multiple anesthesia exposures in and of themselves cause long-term cognitive problems or brain damage. However, other factors such as age, pre-existing medical conditions, and the type and length of surgery may impact cognitive function.

It is important to have an open and honest conversation with your healthcare team about your concerns regarding multiple anesthesia exposures. They can provide you with more information about the risks and benefits of anesthesia and help you make an informed decision about your care.

**Iteration 2** Retrieved on 04/27/2023

It's understandable to have concerns about the potential risks of anesthesia, especially if you are undergoing multiple procedures that require anesthesia. However, the risk of brain damage from anesthesia is extremely low, even with multiple anesthetics.

Modern anesthesia medications and techniques are carefully selected and administered by highly trained professionals to ensure safety and minimize risks. Additionally, anesthesia providers continuously monitor patients' vital signs and adjust medications as needed to maintain appropriate levels of sedation and pain control.

While there is always a small risk of complications associated with anesthesia, including the possibility of adverse effects on the brain, the risk of permanent brain damage from anesthesia is extremely low. In fact, the likelihood of developing brain damage from anesthesia is estimated to be less than 1 in 100,000 procedures.

It's important to discuss any concerns you may have about the risks of anesthesia with your healthcare provider before the procedure. They can provide you with more specific information about the anesthesia process and help alleviate any anxiety you may be experiencing.

**Iteration 3** Retrieved on 04/27/2023

It is normal for patients to have concerns about the potential risks of anesthesia, including the possibility of brain damage. However, the likelihood of experiencing brain damage as a result of anesthesia is extremely rare, particularly when modern anesthesia techniques and equipment are used by trained professionals.

While there is a small risk of cognitive impairment or memory problems associated with anesthesia, particularly in elderly patients or those with pre-existing conditions, the vast majority of patients recover fully without any long-term effects. In fact, many patients report feeling refreshed and rested after waking up from anesthesia.

If a patient requires multiple anesthetics, their healthcare team will take extra precautions to minimize any potential risks and ensure a safe and successful experience. This may include monitoring the patient's vital signs more closely, adjusting the type or dosage of anesthesia used, or providing additional post-operative care and support.

It is important for patients to discuss any concerns they may have with their healthcare provider, including their anesthesiologist, prior to their procedure. This can help to alleviate anxiety and ensure that the patient feels informed and prepared for their anesthesia experience.

Retrieved on 03/14/2023

**Q4. Focus on patients' self-perceptions in preparation for anesthesia. Will I lose my memory?**

It is common for patients to have concerns about the potential effects of anesthesia on memory. However, the vast majority of patients do not experience any long-term memory problems as a result of anesthesia.

While some patients may experience temporary memory loss immediately after the procedure, this is typically short-lived and resolves on its own. The medications used during anesthesia can cause a temporary disruption in the formation of new memories, but this typically does not result in long-term memory problems.

Studies have shown that the risk of long-term memory problems as a result of anesthesia is very low, especially for healthy individuals. However, other factors such as age, pre-existing medical conditions, and the type and length of surgery may impact cognitive function.

It is important to discuss any concerns you have about anesthesia and memory loss with your healthcare team. They can provide you with more information about the risks and benefits of anesthesia and help you make an informed decision about your care. Additionally, they can discuss any strategies or precautions that may help minimize the risk of memory problems during and after anesthesia.

**Iteration 2** Retrieved on 04/27/2023

It's common for patients to have concerns about potential side effects of anesthesia, including memory loss. However, the risk of long-term memory loss from anesthesia is extremely low.

Studies have shown that while some patients may experience short-term memory loss immediately following anesthesia, this typically resolves within a few hours to days. Most patients do not experience any significant long-term effects on memory or cognitive function.

In some cases, patients who are already at higher risk for cognitive impairment or dementia may be more susceptible to postoperative cognitive dysfunction (POCD), which can include memory impairment. However, the risk of POCD is still relatively low, and most patients recover fully within a few weeks to months after surgery.

It's important to discuss any concerns you may have about memory loss or other potential side effects of anesthesia with your healthcare provider before the procedure. They can provide you with more specific information about the anesthesia process and help alleviate any anxiety you may be experiencing. Additionally, it's important to follow all preoperative and postoperative instructions provided by your healthcare team to help ensure a safe and successful procedure.

**Iteration 3** Retrieved on 04/27/2023

It is possible for patients to experience memory loss or confusion following anesthesia, particularly in elderly patients or those with pre-existing cognitive impairments. However, the vast majority of patients do not experience any long-term memory problems as a result of anesthesia.

Temporary memory loss or confusion is more common immediately after waking up from anesthesia, but these effects typically dissipate within a few hours to a few days. In rare cases, patients may experience longer-lasting memory problems or confusion, but these instances are typically associated with pre-existing medical conditions or other factors that increase the risk of cognitive impairment.

It is important for patients to discuss any concerns they may have about memory loss or confusion with their healthcare provider, including their anesthesiologist, prior to their procedure. This can help to alleviate anxiety and ensure that the patient feels informed and prepared for their anesthesia experience.

Additionally, patients can take steps to help minimize the risk of memory problems or confusion following anesthesia. This includes following any instructions provided by their healthcare team, disclosing all medical conditions and medications to their anesthesiologist, and getting plenty of rest and recovery time after their procedure.

Retrieved on 03/14/2023

**Q5. Focus on patients' self-perceptions in preparation for anesthesia. Will I have nausea and vomiting when I wake up?**

Nausea and vomiting are common side effects that some patients experience after waking up from anesthesia. The risk of experiencing these side effects can vary depending on factors such as the type and duration of the surgery, the type of anesthesia used, and the patient's individual health status and susceptibility.

However, there are medications and techniques that can be used to help minimize the risk of nausea and vomiting after anesthesia. Your healthcare team can discuss these options with you and develop a plan to help manage any potential side effects.

In addition, it is important to follow any preoperative instructions provided by your healthcare team to help minimize the risk of nausea and vomiting after anesthesia. These instructions may include avoiding food and drink for a certain period of time before the procedure, adjusting medications as needed, and staying well hydrated.

If you do experience nausea or vomiting after waking up from anesthesia, it is important to let your healthcare team know so they can provide appropriate treatment and support. They may recommend medications to help alleviate these symptoms or other strategies to help you feel more comfortable as you recover.

**Iteration 2** Retrieved on 04/27/2023

Nausea and vomiting are common side effects of anesthesia, especially when patients undergo general anesthesia. However, there are many strategies that can be used to help prevent and manage these symptoms.

Before your procedure, your healthcare provider may recommend that you avoid eating and drinking for a certain period of time to help reduce the risk of nausea and vomiting. Additionally, medications such as anti-nausea drugs may be administered before, during, or after your procedure to help prevent or manage symptoms.

It's important to let your healthcare provider know if you have a history of motion sickness, nausea, or vomiting after previous procedures, as this can increase your risk of experiencing these symptoms again.

If you do experience nausea or vomiting after your procedure, it's important to follow your healthcare provider's instructions for managing these symptoms. This may include taking medications or adjusting your diet and fluid intake.

It's important to discuss any concerns you may have about nausea or vomiting with your healthcare provider before the procedure. They can provide you with more specific information about the anesthesia process and help alleviate any anxiety you may be experiencing. Additionally, following all preoperative and postoperative instructions provided by your healthcare team can help reduce the risk of complications and promote a successful recovery.

**Iteration 3** Retrieved on 04/27/2023

Nausea and vomiting are common side effects of anesthesia, particularly general anesthesia, and may occur when a patient wakes up after a procedure. However, the incidence of post-operative nausea and vomiting (PONV) can be reduced by several methods, and not all patients experience it.

The anesthesiologist and healthcare team can take steps to minimize the risk of nausea and vomiting, such as administering anti-nausea medications before and after the procedure, adjusting the type or dosage of anesthesia used, and providing clear instructions to the patient regarding eating and drinking before the procedure.

Additionally, patients can take steps to help prevent PONV by following any instructions provided by their healthcare team, avoiding eating or drinking anything for several hours prior to the procedure as instructed, staying hydrated before and after the procedure, and taking any anti-nausea medications as prescribed.

If a patient does experience nausea or vomiting after waking up from anesthesia, their healthcare team can provide medication or other treatments to help alleviate these symptoms and ensure their comfort and well-being.

It is important for patients to discuss any concerns or questions they may have about anesthesia and its potential side effects with their healthcare provider, including their anesthesiologist, prior to their procedure. This can help to ensure that the patient feels informed and prepared for their anesthesia experience.

Retrieved on 03/14/2023

**Q6. Focus on patients' self-perceptions in preparation for anesthesia. How does anesthesia work?**

Anesthesia works by using medications to induce a state of unconsciousness or sedation in the patient, allowing for pain-free surgical procedures or other medical interventions. There are several different types of anesthesia, including general anesthesia, regional anesthesia, and local anesthesia, which are chosen based on the type and duration of the procedure and the patient's individual needs.

During general anesthesia, the patient is unconscious and unaware of their surroundings. Medications are administered through an IV or inhalation to induce the state of unconsciousness. The anesthesiologist carefully monitors the patient's vital signs, including heart rate, blood pressure, and oxygen saturation, throughout the procedure to ensure their safety.

Regional anesthesia involves the injection of local anesthetics around a specific area of the body, such as the spinal cord or nerves, to numb the area and provide pain relief during a surgical procedure. The patient remains awake and aware during the procedure, but does not feel any pain or discomfort in the area where the regional anesthesia was administered.

Local anesthesia involves the injection of local anesthetics into a specific area of the body, such as the skin or soft tissue, to provide pain relief during a minor surgical procedure or medical intervention.

Anesthesia works by altering the patient's consciousness and nervous system function, which can have a wide range of effects on the body. However, the medications used during anesthesia are carefully titrated and monitored to achieve the desired level of sedation or unconsciousness while minimizing the risk of complications.

**Iteration 2** Retrieved on 04/27/2023

Anesthesia is a medical treatment used to prevent pain and discomfort during surgery or other medical procedures. There are three main types of anesthesia: general anesthesia, regional anesthesia, and local anesthesia. Each type works in slightly different ways.

General anesthesia involves administering medications that cause a patient to become unconscious and unaware of their surroundings. This is typically done through intravenous (IV) medications or inhaled gases. Once a patient is under general anesthesia, their vital signs and level of consciousness are closely monitored by an anesthesia provider.

Regional anesthesia involves administering medications to numb a specific region of the body, such as an arm or leg. This is typically done through an injection of local anesthetic near the nerves that supply sensation to the affected area. Patients may remain awake and aware during regional anesthesia, or they may be given sedation to help them relax.

Local anesthesia involves administering medications to numb a small area of the body, such as a tooth or a small incision. This is typically done through an injection of local anesthetic at the site of the procedure.

In all cases, anesthesia works by blocking nerve impulses to the brain, which prevents the brain from receiving signals of pain or discomfort. The specific medications and techniques used will depend on the patient's individual needs, medical history, and the type of procedure being performed.

It's important to discuss any questions or concerns you may have about anesthesia with your healthcare provider before the procedure. They can provide you with more specific information about the anesthesia process and help alleviate any anxiety you may be experiencing.

**Iteration 3** Retrieved on 04/27/2023

Anesthesia is a medical treatment that is used to prevent pain and discomfort during medical procedures or surgeries. There are several types of anesthesia, including local anesthesia, regional anesthesia, and general anesthesia. The type of anesthesia used depends on the type of procedure being performed and the patient's individual needs.

General anesthesia is the most common type of anesthesia used for surgeries and more invasive procedures. It works by inducing a state of unconsciousness, in which the patient is completely unaware of their surroundings and does not feel any pain or discomfort. This is typically achieved through the use of intravenous (IV) medications or inhaled gases, which are administered and monitored by an anesthesiologist.

Local anesthesia and regional anesthesia are used for less invasive procedures and work by numbing a specific area of the body. Local anesthesia is typically administered through injection, while regional anesthesia involves numbing larger areas of the body, such as an arm or leg, through injection or a nerve block.

During anesthesia, the anesthesiologist monitors the patient's vital signs, such as heart rate, blood pressure, and oxygen levels, to ensure that the patient remains safe and stable throughout the procedure. After the procedure is complete, the patient is carefully monitored as they wake up from the anesthesia and any side effects or complications are treated as necessary.

It is important for patients to discuss any concerns they may have about anesthesia with their healthcare provider, including their anesthesiologist, prior to their procedure. This can help to ensure that the patient feels informed and prepared for their anesthesia experience.

Retrieved on 03/14/2023

**Q7. Focus on patients' self-perceptions in preparation for anesthesia. What words are most associated with fear of anesthesia?**

Words that are commonly associated with fear of anesthesia include:

- Unconsciousness
- Loss of control
- Needles
- Paralysis
- Death
- Pain
- Nausea
- Vomiting
- Breathing problems
- Memory loss

These words can elicit fear and anxiety in some patients who are preparing for anesthesia. It is important for healthcare providers to be aware of these concerns and provide information and support to help alleviate fears and ensure a safe and comfortable experience for the patient. Open communication between the patient and their healthcare team can help address any questions or concerns and build trust between the patient and the providers.

**Iteration 2** Retrieved on 04/27/2023

Some of the most common words associated with the fear of anesthesia include:

- Loss of control: Many people fear the loss of control that comes with being unconscious or sedated during anesthesia.
- Side effects: Concerns about potential side effects of anesthesia, such as nausea, vomiting, or memory loss, can also contribute to anxiety and fear.
- Pain: Patients may be afraid of experiencing pain or discomfort during or after the procedure, despite the use of anesthesia.
- Death: The possibility of a serious complication or death related to anesthesia can also be a source of fear for some patients.
- Needles: Fear of needles or injections is a common phobia and may contribute to anxiety surrounding anesthesia.
- Unknown: Some patients may simply be afraid of the unknown and uncertainty of what to expect during and after anesthesia.

It's important to discuss any fears or concerns you may have with your healthcare provider before the procedure. They can provide you with more specific information about the anesthesia process and help alleviate any anxiety you may be experiencing. Additionally, many healthcare facilities offer preoperative education programs that can help patients better understand what to expect before, during, and after the procedure.

**Iteration 3** Retrieved on 04/27/2023

The fear of anesthesia, also known as anesthesiophobia, can be a common concern for patients undergoing medical procedures. Some of the words and phrases that are commonly associated with this fear include:

1. Loss of control: Patients may fear that they will lose control of their body or be unable to communicate during the procedure.
2. Not waking up: Patients may be concerned that they will not wake up from the anesthesia or that they will experience complications during the procedure.
3. Memory loss: Patients may worry that they will experience memory loss or confusion following the procedure.
4. Nausea and vomiting: Patients may be concerned about experiencing nausea and vomiting after waking up from the anesthesia.
5. Pain: Patients may worry that they will experience pain during or after the procedure.
6. Breathing difficulties: Patients may be concerned about experiencing breathing difficulties or complications during the procedure.
7. Allergic reactions: Patients may worry about having an allergic reaction to the anesthesia or other medications used during the procedure.

It is important for patients to discuss any concerns they may have about anesthesia with their healthcare provider, including their anesthesiologist, prior to their procedure. This can help to alleviate anxiety and ensure that the patient feels informed and prepared for their anesthesia experience.

Retrieved on 03/14/2023

**Q8 . Focus on patients' self-perceptions in preparation for anesthesia. What does an anesthesiologist do?**

An anesthesiologist is a medical doctor who specializes in administering anesthesia and managing a patient's pain during and after surgical procedures or medical interventions. They work closely with the surgical team and other healthcare providers to ensure the patient's safety and comfort during their procedure.

The role of an anesthesiologist typically involves the following:

- Preoperative evaluation: An anesthesiologist will evaluate the patient's medical history and current health status to determine the most appropriate type of anesthesia for the procedure and minimize any potential risks.
- Administering anesthesia: An anesthesiologist administers anesthesia medications to induce and maintain a state of unconsciousness or sedation during the procedure. They carefully monitor the patient's vital signs, such as heart rate, blood pressure, and oxygen saturation, to ensure their safety throughout the procedure.
- Pain management: An anesthesiologist may administer pain-relieving medications or techniques, such as nerve blocks or epidural anesthesia, to manage a patient's pain during and after the procedure.
- Postoperative care: An anesthesiologist will monitor the patient as they recover from the anesthesia and manage any potential side effects or complications, such as nausea or breathing problems.
- Patient education: An anesthesiologist may provide education and support to the patient and their family about the anesthesia process and what to expect before, during, and after the procedure.

Anesthesiologists play a critical role in ensuring the safety and comfort of patients undergoing surgical procedures or medical interventions that require anesthesia. They work closely with the surgical team and other healthcare providers to provide individualized care and support throughout the patient's entire experience.

**Iteration 2** Retrieved on 04/27/2023

An anesthesiologist is a medical doctor who specializes in administering anesthesia and managing pain relief for patients undergoing surgical or other medical procedures. Their primary responsibility is to ensure the safety and comfort of the patient during the procedure.

Before the procedure, the anesthesiologist will review the patient's medical history, perform a physical exam, and discuss the anesthesia options with the patient. They will also monitor the patient's vital signs during the procedure and adjust the anesthesia as needed to maintain the appropriate level of sedation and pain relief.

During the procedure, the anesthesiologist may use a variety of techniques to administer anesthesia, including intravenous medications, inhaled gases, or nerve blocks. They will also monitor the patient's breathing, heart rate, blood pressure, and other vital signs to ensure that the patient remains safe and comfortable.

After the procedure, the anesthesiologist may continue to monitor the patient's recovery and provide pain management as needed. They may also provide recommendations for managing any side effects of the anesthesia, such as nausea or dizziness.

Overall, the role of an anesthesiologist is critical to the success of a surgical or medical procedure. They work closely with the patient and the surgical team to ensure that the patient is as comfortable and safe as possible before, during, and after the procedure.

**Iteration 3** Retrieved on 04/27/2023

An anesthesiologist is a medical doctor who specializes in administering anesthesia to patients undergoing medical procedures or surgeries. They work closely with the surgical team and other healthcare providers to ensure that patients receive safe and effective anesthesia care.

Before the procedure, the anesthesiologist will evaluate the patient's medical history and current health status to determine the appropriate type and dosage of anesthesia to use. They will also discuss the risks and benefits of the anesthesia with the patient and obtain their consent.

During the procedure, the anesthesiologist administers the anesthesia and monitors the patient's vital signs, such as heart rate, blood pressure, and oxygen levels, to ensure that they remain safe and stable throughout the procedure. They may also adjust the type or dosage of anesthesia as necessary based on the patient's response.

After the procedure, the anesthesiologist carefully monitors the patient as they wake up from the anesthesia and any side effects or complications are treated as necessary.

Anesthesiologists also play an important role in managing pain during and after the procedure. They may administer pain medication or other treatments to help manage pain and ensure the patient's comfort and well-being.

Overall, the anesthesiologist's role is to ensure that the patient receives safe and effective anesthesia care, while minimizing any potential risks or complications. It is important for patients to discuss any questions or concerns they may have about anesthesia with their healthcare provider, including their anesthesiologist, prior to their procedure.

Retrieved on 03/14/2023

**Q9. Focus on patients' self-perceptions in preparation for anesthesia. What is intubation?**

Intubation is a medical procedure where a tube is inserted into a patient's airway through their mouth or nose to help them breathe. This is commonly done when a patient undergoes general anesthesia for surgery. The tube, also known as an endotracheal tube, is connected to a ventilator that delivers oxygen and anesthetic gases to the patient's lungs while also removing carbon dioxide.

During intubation, the patient is usually unconscious due to the effects of anesthesia, and their airway is temporarily blocked to prevent food, saliva, or vomit from entering the lungs. The procedure is typically performed by an anesthesiologist or a trained healthcare professional, who uses specialized instruments such as a laryngoscope to visualize the airway and guide the tube into place.

Intubation is a crucial aspect of anesthesia care and is performed to ensure the patient's safety during surgery. However, it can be a source of anxiety and discomfort for some patients. It is important for healthcare providers to communicate clearly with patients before the procedure, provide appropriate pain management and sedation, and answer any questions or concerns the patient may have to alleviate their anxiety.

**Iteration 2** Retrieved on 04/27/2023

Intubation is a medical procedure in which a flexible tube is inserted through the mouth and into the trachea (windpipe) to help a patient breathe during anesthesia or other medical procedures. The tube is usually inserted while the patient is under general anesthesia to prevent them from experiencing discomfort or breathing difficulties.

During intubation, the anesthesiologist or another healthcare provider will use a laryngoscope to visualize the patient's airway and insert the endotracheal tube (ETT) through the mouth and into the trachea. The ETT is then connected to a machine that delivers oxygen and anesthesia gases, as well as monitors the patient's breathing and vital signs.

Intubation may also be used in emergency situations, such as when a patient is experiencing respiratory distress or cannot breathe on their own. In these cases, the procedure may be done quickly without the use of general anesthesia.

After the procedure, the ETT is typically removed when the patient is awake and able to breathe on their own. Patients may experience a sore throat or coughing as a result of the procedure, but these symptoms typically resolve within a few days.

Intubation is a common procedure in anesthesia and critical care, and is generally considered safe when performed by a qualified healthcare provider. However, as with any medical procedure, there are potential risks and complications that should be discussed with your healthcare provider before the procedure.

**Iteration 3** Retrieved on 04/27/2023

Intubation is a medical procedure that involves the insertion of a tube into the patient's airway in order to maintain an open airway and facilitate breathing. It is often used during general anesthesia, as well as in other medical situations where a patient may have difficulty breathing on their own.

During intubation, the patient is typically sedated or given anesthesia to prevent discomfort. The tube is then inserted through the mouth or nose and down into the trachea, or windpipe. Once in place, the tube is connected to a ventilator or other breathing machine, which helps the patient to breathe.

Intubation is usually performed by a trained healthcare provider, such as an anesthesiologist, respiratory therapist, or emergency medical technician (EMT). The procedure is generally considered safe, but there are some risks, such as damage to the vocal cords, bleeding, or infection.

After the procedure, the patient is typically closely monitored to ensure that the tube remains in the correct position and that there are no complications. Once the patient is no longer in need of the tube, it is removed and the patient is allowed to breathe on their own.

Intubation can be a source of anxiety or concern for some patients, particularly those who have never undergone the procedure before. It is important for patients to discuss any questions or concerns they may have about intubation with their healthcare provider, including their anesthesiologist, prior to their procedure.

Retrieved on 03/14/2023

**Q10. Focus on patients' self-perceptions in preparation for anesthesia. Will I be on a machine that breathes for me? (or something similar)**

Patients who are scheduled to undergo general anesthesia may have concerns and questions about their experience with the anesthesia process. One common question that patients ask is whether they will be on a machine that breathes for them during the procedure.

The answer is yes, during general anesthesia, patients are typically connected to a ventilator that helps them breathe. The ventilator delivers a mixture of oxygen and anesthetic gases to the patient's lungs and removes carbon dioxide, ensuring adequate oxygenation and ventilation. The ventilator is controlled by the anesthesia provider, who monitors the patient's breathing and adjusts the settings as necessary to maintain the patient's vital signs.

It's important for healthcare providers to communicate clearly with patients before the procedure and provide them with information about what to expect during anesthesia. Patients may feel more comfortable if they understand the equipment and processes that will be used to keep them safe and comfortable during the procedure.

Healthcare providers should also address any concerns or fears that the patient may have, provide appropriate pain management and sedation, and answer any questions they may have to alleviate their anxiety. By focusing on patients' self-perceptions in preparation for anesthesia, healthcare providers can help ensure a positive patient experience and promote better outcomes.

**Iteration 2** Retrieved on 04/27/2023

Yes, it is common for patients undergoing general anesthesia to be placed on a mechanical ventilator, which is a machine that assists with breathing during the procedure. The ventilator delivers oxygen and anesthetic gases to the patient's lungs and also removes carbon dioxide from the body.

The use of a mechanical ventilator is necessary because general anesthesia can cause temporary suppression of the respiratory drive, making it difficult for the patient to breathe on their own. The machine helps ensure that the patient receives enough oxygen and is able to maintain adequate ventilation throughout the procedure.

The anesthesiologist or another healthcare provider will monitor the patient's breathing and adjust the ventilator settings as needed to ensure that the patient is comfortable and safe. After the procedure, the patient will typically be weaned off the ventilator gradually as they regain consciousness and their respiratory drive returns.

Although the use of a mechanical ventilator may sound intimidating, it is a routine part of the anesthesia process and is generally considered safe when used appropriately by a qualified healthcare provider. Your healthcare provider can provide you with more specific information about what to expect during the procedure and can address any concerns or questions you may have.

**Iteration 3** Retrieved on 04/27/2023

Yes, it is possible that you may be placed on a machine that breathes for you during the administration of anesthesia. This machine is called a mechanical ventilator, and it is commonly used during general anesthesia and other medical procedures that require the patient to be sedated or unconscious.

The mechanical ventilator helps to maintain the patient's breathing by delivering oxygen and removing carbon dioxide from the lungs. It can also help to regulate the patient's respiratory rate and volume of air moved in and out of the lungs.

The use of a mechanical ventilator can be a source of anxiety or concern for some patients, particularly those who have never undergone anesthesia before. It is important to discuss any questions or concerns you may have with your healthcare provider, including your anesthesiologist, prior to your procedure. They can help to explain the procedure, the use of the mechanical ventilator, and any potential risks or complications that may be associated with its use. They can also help to address any fears or concerns you may have and help you feel more informed and prepared for your anesthesia experience.

Retrieved on 03/14/2023

**Q11. Focus on patients' self-perceptions in preparation for anesthesia. Will I be in pain after surgery?**

Patients who are scheduled to undergo surgery and receive anesthesia may have concerns about pain management after the procedure. One common question that patients ask is whether they will experience pain after surgery.

The answer to this question depends on several factors, including the type of surgery, the patient's overall health, and the type of anesthesia used. However, healthcare providers can take several steps to minimize pain and discomfort after surgery.

Before the procedure, healthcare providers should discuss pain management options with the patient and develop an individualized pain management plan that takes into account the patient's medical history and personal preferences. This may include the use of medications, such as opioids or non-opioid pain relievers, as well as non-pharmacological pain management techniques, such as relaxation exercises or physical therapy.

After the procedure, healthcare providers should monitor the patient's pain levels and adjust the pain management plan as needed. Patients should be encouraged to communicate any pain or discomfort they experience, and healthcare providers should provide education and support to help them manage their pain effectively.

It's important to note that some discomfort or pain after surgery is normal and expected, and patients should be prepared for this possibility. However, with appropriate pain management, most patients can achieve a comfortable level of pain control and recover more smoothly.

By focusing on patients' self-perceptions in preparation for anesthesia and addressing their concerns about pain management after surgery, healthcare providers can help ensure a positive patient experience and promote better outcomes.

**Iteration 2** Retrieved on 04/27/2023

It is common to experience some pain or discomfort after surgery, but the amount and duration of pain can vary depending on the type of surgery and the individual patient's response to the procedure.

Before the surgery, the anesthesiologist or another healthcare provider will discuss pain management options with you, including medications that can be used to control pain both during and after the procedure. The goal is to manage your pain effectively so that you are comfortable and can begin to resume normal activities as soon as possible.

There are several types of pain management medications that may be used after surgery, including opioids, nonsteroidal anti-inflammatory drugs (NSAIDs), and local anesthetics. The specific medication or combination of medications used will depend on a variety of factors, including the type of surgery, the patient's medical history, and any allergies or other factors that may affect the patient's response to the medication.

It is important to follow the instructions of your healthcare provider regarding pain management after surgery, including the proper dosing and timing of medications. You may also be given instructions for managing any side effects of the medications, such as nausea or dizziness.

If you experience significant or prolonged pain after surgery, it is important to contact your healthcare provider so that they can adjust your pain management plan or investigate potential underlying causes of the pain.

**Iteration 3** Retrieved on 04/27/2023

It is common to experience some degree of pain or discomfort after surgery. However, the severity and duration of pain can vary depending on the type of surgery, the individual patient's pain tolerance, and other factors.

To help manage pain after surgery, your healthcare team will typically use a combination of medications and other pain management techniques. These may include:

- Pain medications, such as opioids or non-opioid pain relievers, which can be given orally, through an IV, or as an injection
- Local anesthetics, which are injected near the surgical site to numb the area and provide pain relief
- Regional anesthesia, which involves blocking pain signals to a larger area of the body, such as an arm or leg, using an injection or nerve block
- Non-pharmacologic pain management techniques, such as relaxation exercises, distraction techniques, or acupuncture
- It is important to discuss any concerns you may have about pain management with your healthcare team, including your anesthesiologist and surgeon, prior to your procedure. They can help to develop a plan for managing pain after surgery that is tailored to your individual needs and preferences.

Additionally, it is important to follow any post-operative instructions provided by your healthcare team, such as taking pain medications as prescribed, keeping the surgical site clean and dry, and avoiding certain activities or movements that may exacerbate pain or delay healing. By working closely with your healthcare team and following their recommendations, you can help to manage pain after surgery and ensure a smoother recovery.

#### S2 (c) Bard responses to the queries listed in Table 1 (Q.1-11)

**Q1. Focus on patients' self-perceptions preparation for anesthesia. What are the most commonly asked question by patients regarding anesthesia?**

**Draft 1**

The most commonly asked question by patients regarding anesthesia is "Will I wake up during surgery?" This is a valid concern, as patients are often anxious about the thought of being awake during surgery and experiencing pain. However, the risk of waking up during surgery is extremely low, thanks to modern advances in anesthesia.

Another common question is "Will I feel any pain?" Patients are often worried about feeling pain during surgery, even if they are unconscious. However, anesthesia is designed to block pain, so patients should not experience any pain during surgery.

Patients may also ask about the side effects of anesthesia. The most common side effects of anesthesia are nausea, vomiting, and dizziness. These side effects are usually temporary and go away within a few hours of the surgery.

Finally, patients may ask about the risks of anesthesia. The risks of anesthesia are very low, but they do exist. Some of the risks of anesthesia include allergic reactions, bleeding, and infection. However, the benefits of anesthesia far outweigh the risks.

If you have any questions about anesthesia, please talk to your doctor or anesthesiologist. They will be able to answer your questions and help you feel more comfortable about your upcoming surgery.

**Draft 2**

Here are the most commonly asked questions by patients regarding anesthesia:

1. **What are the risks of anesthesia?**

The risks of anesthesia are very low, but they can include allergic reactions, problems with breathing, and heart problems. It is important to discuss your medical history with your doctor before having surgery so that they can assess your risk.

2. **How long will I be unconscious?**

The length of time you will be unconscious depends on the type of anesthesia you receive and the length of your surgery. You may be awake during some procedures, but you will not feel any pain.

3. **Will I feel any pain?**

You will not feel any pain during surgery or immediately afterwards. However, you may experience some pain after your surgery. Your doctor will prescribe pain medication to help you manage the pain.

4. **What will happen after I wake up?**

Once you wake up after surgery, you will be in a recovery room. You will be monitored by nurses and doctors. You may also be given some fluids and medications. You will be able to go home once you are able to eat and drink on your own.

5. **What should I do to prepare for anesthesia?**

Here are some things you can do to prepare for anesthesia:

- Tell your doctor about all of your medications, including prescription and over-the-counter medications.
- Tell your doctor about any allergies you have.
- Stop smoking and drinking alcohol at least 24 hours before your surgery.
- Eat a light breakfast on the day of your surgery.

It is important to follow your doctor's instructions carefully before and after your surgery.

**Draft 3**

The most commonly asked questions by patients regarding anesthesia are:

- What is the chance of a serious complication from anesthesia?
- Why is it important not to eat or drink anything after midnight?
- What will I feel during the surgery?
- Will I wake up during the surgery?
- How long will it take to wake up after the surgery?
- Will I have any pain after the surgery?
- How long will it take to recover from the surgery?
- When can I go home?
- Can I drive after the surgery?

It is important to answer these questions honestly and to provide reassurance to the patient. The patient should also be encouraged to ask any other questions that they may have.

Here are some tips for preparing patients for anesthesia:

- Explain the risks and benefits of anesthesia.
- Answer any questions that the patient may have.
- Reassure the patient that they will be safe.
- Encourage the patient to ask any other questions that they may have.

Here are some things that patients can do to prepare for anesthesia:

- Follow the instructions of their doctor or nurse.
- Avoid eating or drinking anything after midnight.
- Take any prescribed medications as directed.
- Get a good night's sleep.
- Relax and try to stay calm.

**Q2. Focus on patients' self-perceptions preparation for anesthesia. Will I ever wake up again?**

Yes, you will wake up again after anesthesia. Anesthesia is a safe and effective way to manage pain and allow surgery to take place. The vast majority of people who have anesthesia wake up without any problems.

The risk of not waking up from anesthesia is very small. In fact, it is estimated that the risk of death from anesthesia is about 1 in 250,000. This means that you are more likely to die from being struck by lightning than from anesthesia.

If you are concerned about waking up from anesthesia, you should talk to your doctor or nurse. They can answer any questions you have and help you feel more comfortable.

**Draft 2**

Yes, you will wake up again after anesthesia. Anesthesia is a safe and effective way to numb the pain during surgery. It does not cause you to lose consciousness permanently. In fact, most people wake up feeling refreshed and ready to go home.

The amount of time it takes to wake up after anesthesia depends on the type of anesthesia you receive and the length of your surgery. Most people wake up within a few hours of the end of their surgery. However, some people may take a few days to fully wake up.

**Draft 3**

Yes, you will wake up again after anesthesia. Anesthesia is a safe and effective way to reduce pain and anxiety during surgery. It works by making you unconscious so that you don't feel any pain. You will wake up when the anesthesia wears off, usually within a few hours of the surgery.

It is important to remember that anesthesia is a temporary state. You will not lose consciousness forever. You will wake up and be able to go home and resume your normal activities.

If you are concerned about anesthesia, you should talk to your doctor or nurse. They can answer any questions that you have and help you feel more comfortable about the procedure.

**Q3. Focus on patients' self-perceptions in preparation for anesthesia. If I have multiple anesthetics, will I get brain damage?**

The risk of brain damage from anesthesia is very low. In fact, it is estimated that only about 1 in 100,000 people who receive general anesthesia will experience any permanent brain damage.

The risk of brain damage from anesthesia is higher in people who have certain medical conditions, such as a history of stroke or heart attack. However, even in these people, the risk is still very low.

There are a few things that you can do to reduce the risk of brain damage from anesthesia:

- Make sure that your doctor knows about all of your medical conditions.
- Tell your doctor about any medications that you are taking.
- Get a good night's sleep before your surgery.
- Relax and try to stay calm before your surgery.

If you are concerned about the risk of brain damage from anesthesia, you should talk to your doctor. They can answer any questions that you have and help you feel more comfortable about the procedure.

**Draft 2**

The short answer is no, you will not get brain damage from having multiple anesthetics.

Anesthesia is a safe and effective way to reduce pain and anxiety during surgery. It works by making you unconscious so that you don't feel any pain. You will wake up when the anesthesia wears off, usually within a few hours of the surgery.

Anesthesia is a temporary state. You will not lose consciousness forever. You will wake up and be able to go home and resume your normal activities.

There is no evidence that having multiple anesthetics causes brain damage. In fact, studies have shown that anesthesia does not have any long-term effects on the brain.

If you are concerned about anesthesia, you should talk to your doctor or nurse. They can answer any questions that you have and help you feel more comfortable about the procedure.

**Draft 3**

The risk of brain damage from multiple anesthetics is extremely small. In fact, the risk of brain damage from a single anesthetic is also very small. Anesthesia is a safe and effective way to reduce pain and anxiety during surgery. It works by making you unconscious so that you don't feel any pain.

There is no evidence that multiple anesthetics increase the risk of brain damage. In fact, some studies have shown that multiple anesthetics may actually reduce the risk of brain damage. This is because multiple anesthetics can help to protect the brain from damage caused by lack of oxygen or blood flow.

If you are concerned about the risk of brain damage from anesthesia, you should talk to your doctor or nurse. They can answer any questions that you have and help you feel more comfortable about the procedure.

Here are some things that you can do to reduce the risk of brain damage from anesthesia:

- Choose a qualified and experienced anesthesiologist.
- Make sure that you are healthy and have no underlying medical conditions.
- Follow the instructions of your doctor or nurse.
- Ask any questions that you have about anesthesia.

**Q4. Focus on patients' self-perceptions in preparation for anesthesia. Will I lose my memory?**

The risk of losing your memory after anesthesia is very small. In fact, most people do not experience any problems with memory after anesthesia. However, some people may experience temporary memory problems, such as difficulty remembering recent events or names. These problems usually go away within a few days or weeks.

There are a few things that you can do to reduce the risk of memory problems after anesthesia:

- Get a good night's sleep before your surgery.
- Avoid alcohol and drugs before your surgery.
- Tell your doctor about any medications that you are taking.
- Talk to your doctor about your concerns about memory problems.

If you are concerned about memory problems after anesthesia, you should talk to your doctor. They can help you to understand the risks and benefits of anesthesia and can recommend ways to reduce the risk of memory problems.

**Draft 2**

It is unlikely that you will lose your memory after anesthesia. However, some people may experience temporary memory problems, such as difficulty remembering recent events or difficulty concentrating. These problems usually go away within a few days or weeks.

If you are concerned about the risk of memory problems after anesthesia, you should talk to your doctor or nurse. They can answer any questions that you have and help you feel more comfortable about the procedure.

Here are some things that you can do to reduce the risk of memory problems after anesthesia:

- Get a good night's sleep before your surgery.
- Avoid alcohol and caffeine in the days leading up to your surgery.
- Talk to your doctor or nurse about any medications that you are taking.
- Ask any questions that you have about anesthesia.

**Draft 3**

The risk of losing your memory after anesthesia is very small. Most people who undergo anesthesia do not experience any problems with their memory. However, some people may experience temporary memory problems, such as difficulty remembering recent events or names. These problems usually go away within a few days or weeks.

There are a few things that you can do to reduce the risk of memory problems after anesthesia:

- Choose a qualified and experienced anesthesiologist.
- Make sure that you are healthy and have no underlying medical conditions.
- Follow the instructions of your doctor or nurse.
- Ask any questions that you have about anesthesia.
- Get a good night's sleep before your surgery.
- Relax and try to stay calm.

If you are concerned about memory problems after anesthesia, you should talk to your doctor or nurse. They can answer any questions that you have and help you feel more comfortable about the procedure.

**Q5. Focus on patients' self-perceptions in preparation for anesthesia. Will I have nausea and vomiting when I wake up?**

Nausea and vomiting after anesthesia are common, but they usually go away quickly. There are a few things that you can do to reduce the risk of nausea and vomiting after anesthesia:

- Tell your doctor or nurse if you are prone to nausea and vomiting.
- Avoid eating or drinking anything after midnight before your surgery.
- Take any prescribed medications as directed.
- Get a good night's sleep before your surgery.
- Relax and try to stay calm.

If you do experience nausea and vomiting after anesthesia, there are a few things that you can do to help relieve the symptoms:

- Sit up or stand up.
- Breathe deeply.
- Use a cool compress on your forehead.
- Sip on ginger ale or water.
- Take over-the-counter medications, such as Dramamine or Phenergan.

If your nausea and vomiting are severe, or if they do not improve after a few hours, you should contact your doctor or nurse.

**Draft 2**

Nausea and vomiting are common side effects of anesthesia, especially in children. However, they can also occur in adults. The risk of nausea and vomiting is higher in people who have a history of these problems, or who are having surgery on their stomach or intestines.

There are a few things that you can do to reduce the risk of nausea and vomiting after anesthesia:

- Tell your doctor or nurse about any history of nausea or vomiting.
- Take any prescribed medications as directed.
- Get a good night's sleep before your surgery.
- Relax and try to stay calm.
- Avoid eating or drinking anything after midnight.
- Ask your doctor or nurse about using anti-nausea medications.

If you do experience nausea and vomiting after anesthesia, there are a few things that you can do to relieve your symptoms:

- Sit upright and lean forward.
- Breathe deeply.
- Suck on ice chips or frozen fruit.
- Drink clear liquids, such as water or ginger ale.
- Take over-the-counter anti-nausea medications, such as Dramamine or Gravol.
- Ask your doctor or nurse about using prescription anti-nausea medications.

Nausea and vomiting after anesthesia are usually temporary and go away within a few hours. However, if your symptoms are severe or do not go away, you should talk to your doctor or nurse.

**Draft 3**

Nausea and vomiting are common after anesthesia. In fact, about 20-30% of people who undergo anesthesia experience nausea and vomiting. The risk of nausea and vomiting is higher in children and people who have motion sickness. There are a few things that you can do to reduce the risk of nausea and vomiting after anesthesia:

- Talk to your doctor or nurse about taking anti-nausea medication before your surgery.
- Avoid eating or drinking anything after midnight before your surgery.
- Stay hydrated by drinking plenty of fluids before and after your surgery.
- Get a good night's sleep before your surgery.
- Relax and try to stay calm before and after your surgery.

If you do experience nausea and vomiting after anesthesia, there are a few things that you can do to feel better:

- Drink plenty of fluids.
- Take over-the-counter anti-nausea medication.
- Avoid eating or drinking anything until you feel better.
- Rest and relax.

**Q6. Focus on patients' self-perceptions in preparation for anesthesia. How does anesthesia work?**

Anesthesia is a state of unconsciousness that is induced by drugs. It is used to reduce pain and anxiety during surgery. The drugs used to induce anesthesia work by blocking signals in the nervous system. This prevents the brain from receiving signals from the body, which causes the patient to lose consciousness.

There are two main types of anesthesia: general anesthesia and regional anesthesia. General anesthesia is used for surgeries that require the patient to be unconscious. Regional anesthesia is used for surgeries that only require the patient to be numb in a specific area of the body.

General anesthesia is usually given by intravenous injection or inhalation. The drugs are absorbed into the bloodstream and travel to the brain, where they block signals in the nervous system. The patient loses consciousness within a few minutes of the drugs being administered.

Regional anesthesia is usually given by injection into the spinal cord or nerves. The drugs block signals in the nervous system in a specific area of the body. The patient remains conscious, but they will not feel pain in the area that is numb.

Anesthesia is a safe and effective way to reduce pain and anxiety during surgery. However, there are some risks associated with anesthesia. The most common risks are:

- Allergic reaction to the drugs
- Side effects of the drugs
- Problems with the airway
- Problems with the heart

The risks of anesthesia are higher in people who have certain medical conditions, such as heart disease, lung disease, or liver disease. It is important to talk to your doctor about the risks of anesthesia before you have any surgery.

**Draft 2**

Anesthesia is a state of unconsciousness produced by a drug or combination of drugs. It is used to prevent pain during surgery and other medical procedures. There are two main types of anesthesia: general anesthesia and local anesthesia.

General anesthesia is a state of unconsciousness that affects the entire body. It is used for major surgeries and other medical procedures that require the patient to be unconscious. General anesthesia is given by intravenous injection or inhalation.

Local anesthesia is a state of unconsciousness that affects a specific area of the body. It is used for minor surgeries and other medical procedures that do not require the patient to be unconscious. Local anesthesia is injected into the area that will be numbed.

Anesthesia works by blocking the transmission of nerve signals. This prevents the patient from feeling pain. Anesthesia also affects the brain, causing the patient to lose consciousness.

Anesthesia is a safe and effective way to reduce pain and anxiety during surgery and other medical procedures. However, there are some risks associated with anesthesia. The most common risk is nausea and vomiting. Other risks include allergic reactions, infection, and breathing problems.

If you are considering anesthesia, it is important to talk to your doctor or nurse about the risks and benefits. They can help you decide if anesthesia is right for you.

**Draft 3**

Anesthesia is a state of unconsciousness created by a combination of drugs. It is used during surgery to prevent pain and anxiety. Anesthesia also helps to relax the muscles and make it easier for the surgeon to operate.

There are two main types of anesthesia: general anesthesia and local anesthesia.

- General anesthesia is a state of unconsciousness that is induced by a combination of drugs. The drugs are injected into a vein or breathed in through a mask. General anesthesia causes a loss of consciousness, pain relief, and muscle relaxation.
- Local anesthesia is a type of anesthesia that numbs a specific area of the body. The drugs are injected into the skin or a nerve. Local anesthesia does not cause a loss of consciousness, but it does cause pain relief and muscle relaxation in the area that is numbed.

Anesthesia works by blocking the signals that are sent from the brain to the body. These signals are responsible for pain, anxiety, and muscle movement. When these signals are blocked, the patient does not feel pain, anxiety, or muscle movement.

Anesthesia is a safe and effective way to reduce pain and anxiety during surgery. It is used millions of times each year without any serious problems.

**Q7. Focus on patients' self-perceptions in preparation for anesthesia. What words are most associated with fear of anesthesia?**

The most common words associated with fear of anesthesia are:

- Fear
- Anxiety
- Worry
- Stress
- Discomfort
- Pain
- Loss of control
- Death

Fear of anesthesia is a common and understandable reaction to the prospect of being put to sleep and having surgery. However, it is important to remember that anesthesia is a very safe procedure, and the risks of serious complications are very low.

If you are feeling anxious about anesthesia, there are a number of things you can do to help reduce your fear, including:

- Talk to your doctor or nurse about your fears and concerns. They can help you to understand the risks and benefits of anesthesia and can provide you with information about how to cope with your anxiety.
- Ask your doctor or nurse about the type of anesthesia that will be used during your surgery. There are a number of different types of anesthesia, and each one has its own risks and benefits.
- Ask your doctor or nurse about the risks of anesthesia. They can help you to understand the very low risks of serious complications.
- Ask your doctor or nurse about the benefits of anesthesia. They can help you to understand that anesthesia can help you to have a safe and comfortable surgery.
- Relaxation techniques, such as deep breathing or meditation, can help to reduce anxiety.
- Listening to music or watching a funny movie can help to take your mind off of your fears.
- Spending time with loved ones can help to provide you with support and comfort.

If you are still feeling anxious about anesthesia, you may want to consider talking to a therapist or counselor. They can help you to develop coping mechanisms and strategies for dealing with your fear.

**Draft 2**

Here are some words that are most associated with fear of anesthesia:

- Death
- Loss of control
- Pain
- Anxiety

These words are often used by people who are afraid of anesthesia to describe their fears. For example, someone might say that they are afraid of dying under anesthesia, or that they are afraid of losing control during the procedure.

It is important to remember that these fears are common and that there are many things that can be done to help people who are afraid of anesthesia. Talking to a doctor or nurse about your fears can be a helpful first step. They can answer any questions that you have about anesthesia and can help you to develop a plan to manage your fears.

**Draft 3**

Here are some words that are most associated with fear of anesthesia:

- Death
- Loss of control
- Pain
- Anxiety
- Nausea and vomiting
- Fear of the unknown

It is important to remember that these are just words. They do not necessarily mean that you will experience any of these things if you have anesthesia. In fact, most people who have anesthesia do not experience any of these things.

If you are feeling anxious about anesthesia, you should talk to your doctor or nurse. They can help you to understand the procedure and to develop a plan to manage your anxiety.

**Q8. Focus on patients' self-perceptions in preparation for anesthesia. What does an anesthesiologist do?**

An anesthesiologist is a medical doctor who specializes in the administration of anesthesia. They are responsible for the patient's safety during surgery and for managing their pain and anxiety. Anesthesiologists also work with the surgeon to ensure that the patient is comfortable and that the surgery is successful.

Here are some of the things that an anesthesiologist does:

- Evaluate the patient's health and history to determine the best type of anesthesia for their surgery.
- Administer the anesthesia, monitoring the patient's vital signs and making sure that they are comfortable.
- Manage any complications that may arise during surgery.
- Help the patient to recover from the surgery.

Anesthesiologists are highly trained and experienced professionals who play an important role in the success of surgery. They are responsible for the patient's safety and comfort, and they work closely with the surgeon to ensure that the surgery is a success.

**Draft 2**

An anesthesiologist is a medical doctor who specializes in the care of patients who are undergoing anesthesia. They are responsible for assessing the patient's medical history, current medications, and overall health. They also choose the appropriate type of anesthesia for the patient and monitor the patient's vital signs during the surgery.

Anesthesiologists are also responsible for educating patients about anesthesia and answering any questions that they may have. They work closely with surgeons and other medical professionals to ensure that patients have a safe and comfortable experience during surgery.

Here are some of the tasks that an anesthesiologist does:

- Assess the patient's medical history, current medications, and overall health.
- Choose the appropriate type of anesthesia for the patient.
- Monitor the patient's vital signs during the surgery.
- Educate the patient about anesthesia and answer any questions that they may have.
- Work closely with surgeons and other medical professionals to ensure that patients have a safe and comfortable experience during surgery.

Anesthesiologists are highly trained and experienced medical professionals. They play an important role in ensuring that patients have a safe and comfortable experience during surgery.

**Draft 3**

An anesthesiologist is a medical doctor who specializes in the care of patients before, during, and after surgery. They are responsible for the safe and effective administration of anesthesia, as well as the management of any medical problems that may arise during surgery.

Anesthesiologists work closely with surgeons, nurses, and other healthcare professionals to ensure that patients have a safe and comfortable surgical experience. They also provide pain management and other supportive care to patients after surgery.

Anesthesiologists must have a strong understanding of the human body and how it works. They must also be able to think critically and make quick decisions in emergency situations. In addition, they must be able to communicate effectively with patients and their families, as well as with other healthcare professionals.

If you are considering a career in anesthesia, you should be prepared to undergo rigorous training and education. After completing medical school, you will need to complete a residency program in anesthesia. This will give you the opportunity to learn the skills and knowledge that you need to be a successful anesthesiologist.

**Q9. Focus on patients' self-perceptions in preparation for anesthesia. What is intubation?**

Intubation is a procedure that involves inserting a tube into the airway to help a person breathe. It is commonly used during surgery, but it can also be used in other situations, such as when a person is unable to breathe on their own.

The tube is usually inserted through the mouth or nose and into the trachea (windpipe). Once the tube is in place, it can be connected to a ventilator, which helps to move air in and out of the lungs.

Intubation is a safe and effective procedure, but it can be uncomfortable and there is a risk of complications. The most common complications are:

- Bleeding
- Infection
- Injury to the vocal cords
- Problems with the tube

If you are considering intubation, it is important to talk to your doctor about the risks and benefits.

**Draft 2**

Intubation is a procedure that is used to place a tube into the trachea (windpipe) to help a patient breathe. It is often necessary during surgery, when a patient is unconscious or when their breathing is compromised.

The procedure is usually performed by an anesthesiologist or a nurse anesthetist. They will first insert a laryngoscope, a device that helps to visualize the vocal cords. The tube is then inserted into the trachea and secured in place.

Intubation can be a life-saving procedure, but it does come with some risks. These risks include:

- Injury to the vocal cords or trachea
- Infection
- Difficulty removing the tube
- Allergic reaction to the tube

If you are considering intubation, it is important to discuss the risks and benefits with your doctor.

**Draft 3**

Intubation is a medical procedure that involves inserting a tube into the airway to keep it open and allow for the delivery of oxygen and ventilation. It is commonly used during surgery, when the patient is unconscious and cannot breathe on their own.

Intubation is a safe and effective procedure, but it can be uncomfortable for the patient. It is important to explain the procedure to the patient and answer any questions that they may have.

The procedure is usually performed by an anesthesiologist or a nurse anesthetist. They will first insert a laryngoscope into the patient's mouth to visualize the vocal cords. Then, they will insert the endotracheal tube into the trachea. The tube is secured in place with tape or a strap.

The patient will be able to breathe through the tube and will be ventilated with a machine. They will also be given oxygen through the tube.

Intubation is a necessary procedure for many patients, but it is important to be aware of the risks and discomfort involved.

**Q10. Focus on patients' self-perceptions in preparation for anesthesia. Will I be on a machine that breathes for me? (or something similar)**

Yes, you will be on a machine that breathes for you if you are unconscious during surgery. This is called mechanical ventilation. The machine will deliver oxygen to your lungs and remove carbon dioxide from your blood. You will not be able to breathe on your own while you are on the machine.

Mechanical ventilation is a safe and effective way to keep you alive during surgery. It is also used in other situations, such as when you are critically ill or have a respiratory problem.

If you are concerned about mechanical ventilation, you should talk to your doctor or nurse. They can answer any questions that you have and help you feel more comfortable about the procedure.

**Draft 2**

Yes, you will be on a machine that breathes for you during anesthesia. This is called mechanical ventilation. It is a safe and effective way to ensure that you get enough oxygen during surgery.

The machine will deliver a continuous stream of air and oxygen into your lungs. The machine will also monitor your breathing and make sure that you are getting enough oxygen.

If you are concerned about mechanical ventilation, you should talk to your doctor or nurse. They can answer any questions that you have and help you feel more comfortable about the procedure.

**Draft 3**

Yes, you will be on a machine that breathes for you if you are under general anesthesia. The machine is called an anesthesia machine, and it is used to deliver the anesthesia drugs and to ventilate your lungs.

The anesthesia machine will deliver the drugs through a mask or an IV line. The drugs will make you unconscious and will prevent you from feeling pain. The machine will also ventilate your lungs by pumping air in and out of your lungs. This will help you to breathe and to get the oxygen that you need.

The anesthesia machine is a safe and effective way to deliver anesthesia. It is used millions of times each year without any serious problems.

If you are concerned about the anesthesia machine, you should talk to your doctor or nurse. They can answer any questions that you have and help you to feel more comfortable about the procedure.

**Q11. Focus on patients' self-perceptions in preparation for anesthesia. Will I be in pain after surgery?**

Most patients experience some pain after surgery. The type and severity of pain will depend on the type of surgery, the patient's individual pain tolerance, and the effectiveness of pain management.

There are a number of ways to manage pain after surgery, including:

- Medications, such as opioids and non-steroidal anti-inflammatory drugs (NSAIDs)
- Nerve blocks
- Physical therapy
- Heat or cold therapy
- Massage

It is important to talk to your doctor about your pain management options before surgery. They can help you to choose the best option for you and to develop a plan to manage your pain after surgery.

It is also important to remember that pain is a normal part of the healing process after surgery. It is important to take your medications as prescribed and to follow your doctor's instructions. With proper pain management, you will be able to recover from surgery and get back to your normal activities.

**Draft 2**

There is no definitive answer to this question as it depends on a number of factors, including the type of surgery you are having, your individual pain tolerance, and the effectiveness of the pain medication you receive. However, most people do experience some pain after surgery, even with the use of pain medication.

The type of surgery you are having will determine the location and severity of the pain you experience. For example, if you are having a major surgery, such as open heart surgery, you will likely experience more pain than if you are having a minor surgery, such as a laparoscopic procedure.

Your individual pain tolerance will also affect how much pain you experience. Some people have a higher pain tolerance than others and may be able to tolerate more pain without experiencing any discomfort.

The effectiveness of the pain medication you receive will also affect how much pain you experience. If you receive effective pain medication, you will likely experience less pain than if you do not receive any pain medication.

There are a number of things you can do to help manage the pain after surgery, including:

- Taking over-the-counter pain medication, such as ibuprofen or acetaminophen.
- Using ice or heat therapy.
- Resting.
- Relaxing.

If you are experiencing severe pain after surgery, you should contact your doctor or nurse. They may be able to prescribe stronger pain medication or recommend other ways to manage the pain.

**Draft 3**

Most patients experience some pain after surgery. The amount of pain you experience will depend on the type of surgery you had, the severity of your injury, and your individual pain tolerance.

There are a number of things that can be done to reduce pain after surgery, including:

- Taking pain medication as prescribed
- Applying ice or heat to the area
- Resting the area
- Elevating the area

If you are experiencing severe pain after surgery, you should contact your surgeon or nurse. They may be able to prescribe stronger pain medication or recommend other treatments to help reduce your pain.

It is important to remember that pain is a subjective experience. What one person considers to be a mild pain may be considered a severe pain by another person. There is no right or wrong way to experience pain.

If you are feeling anxious about pain after surgery, you should talk to your doctor or nurse. They can help you to develop a plan to manage your pain and to cope with any anxiety that you may be feeling.

### S3. Exemplary response generated by ChatGPT and Bard

#### S3(a). Exemplary response generated by ChatGPT

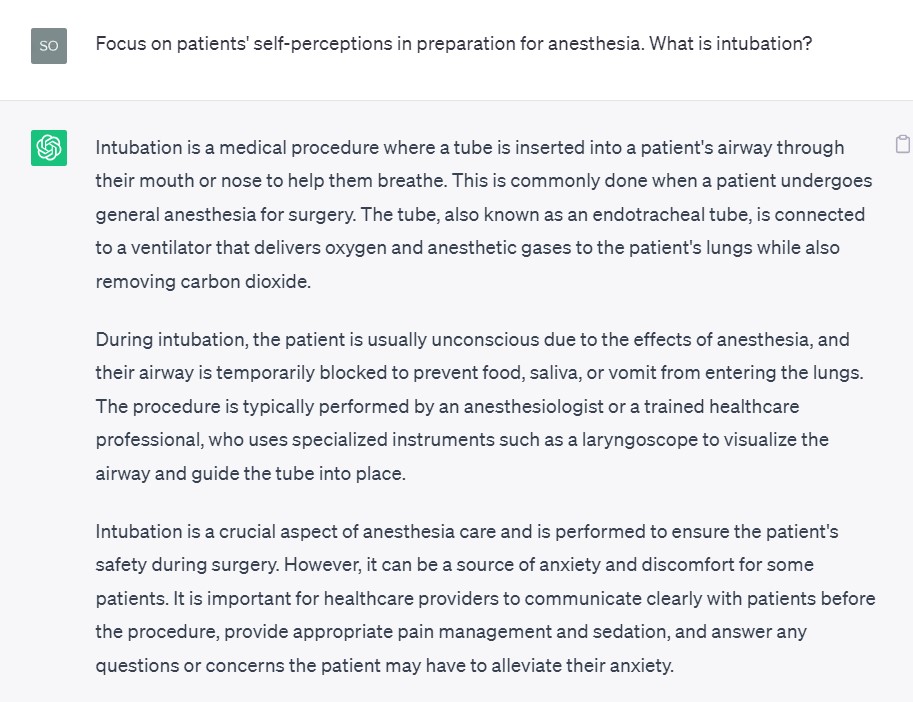

##### Fig. S3(a).1 Example of ChatGPT response to the questions utilized in this study.

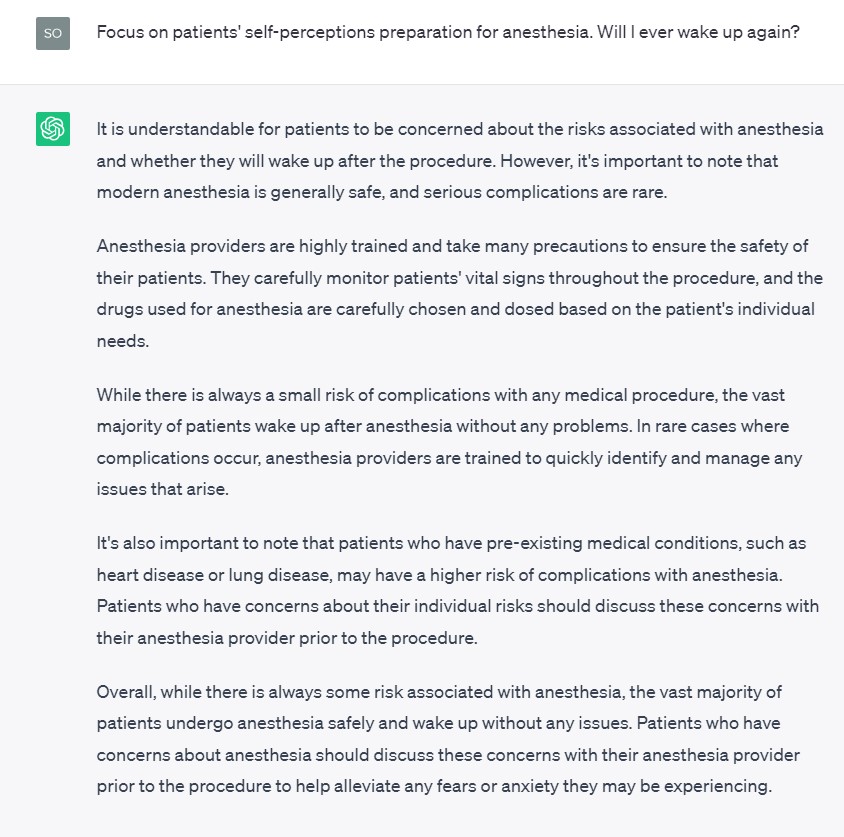

##### Fig. S3(a).2 Example of ChatGPT response to the questions utilized in this study.

#### S3(b). Exemplary response generated by Bard. Errors in responses are highlighted in red text.

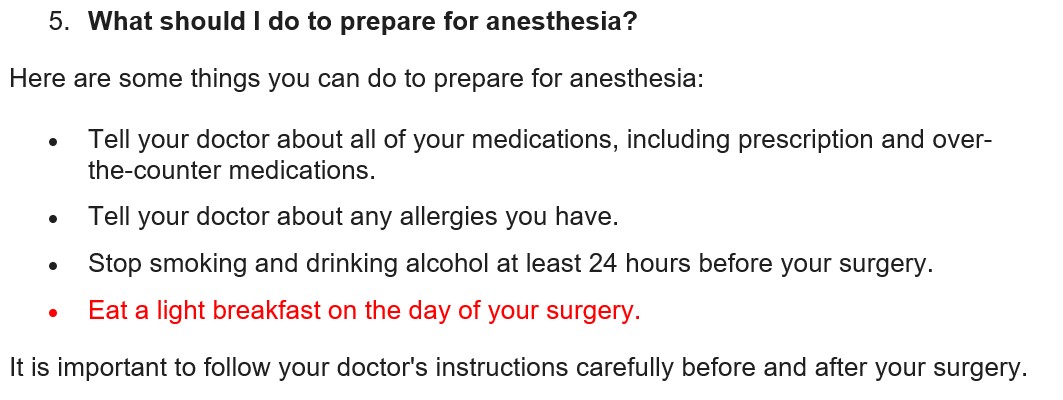

##### Fig. S3(b).1 Exemplary error 1 from Bard.

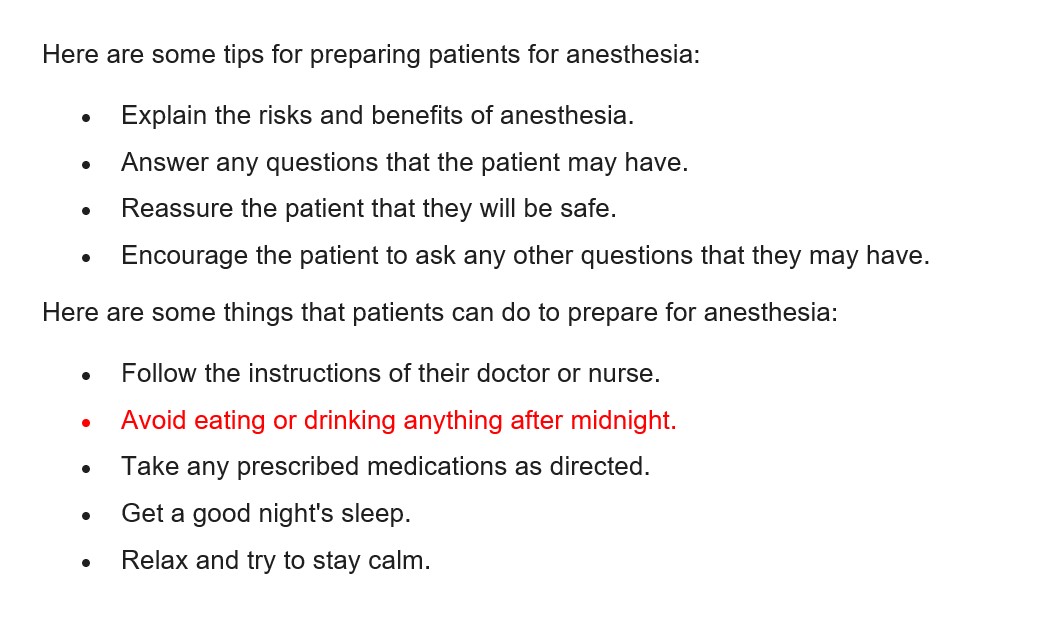

##### Fig. S3(b).2 Exemplary error 2 from Bard.

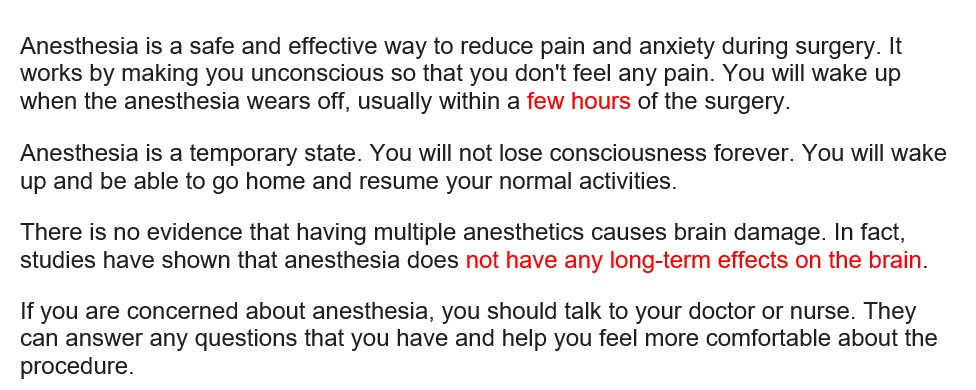

##### Fig. S3(b).3 Exemplary error 3 from Bard.

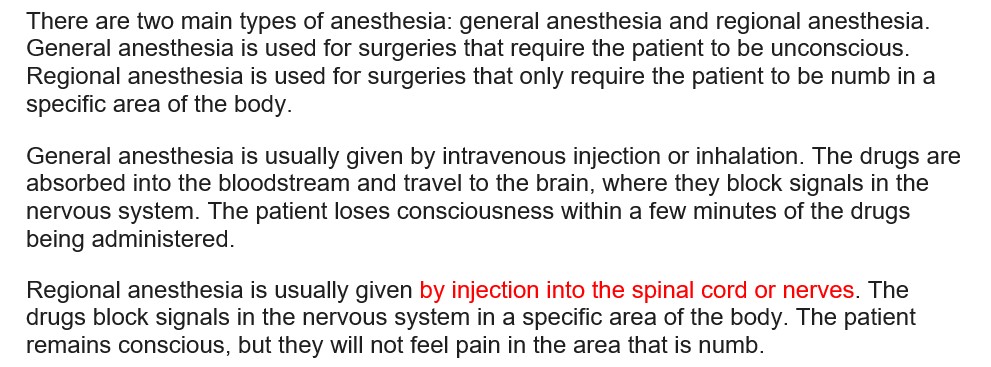

##### Fig. S3(b).4 Exemplary error 4 from Bard.

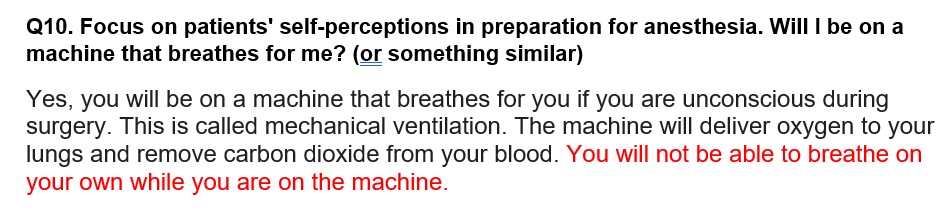

##### Fig. S3(b).5 Exemplary error 5 from Bard.
